# Methylphenidate-induced changes in target-enriched functional connectivity in ADHD

**DOI:** 10.1101/2025.11.26.25341066

**Authors:** Z. van der Pal, L. Reneman, H.M. Geurts, D.E. Boucherie, L. Douw, A. Schrantee

## Abstract

Methylphenidate is the most widely used pharmacological treatment for attention-deficit/hyperactivity disorder (ADHD) and acts primarily by blocking dopamine and noradrenaline transporters, increasing catecholamine availability. However, how these molecular actions translate into large-scale functional brain network changes, and how such changes relate to cognition, remains unclear. Moreover, methylphenidate effects have been shown to depend on age, yet whether this extends to molecularly informed brain networks is unknown. In a pre–post challenge design, 83 stimulant treatment-naïve children and adults with ADHD underwent resting-state fMRI before and after a single methylphenidate dose (0.5 mg/kg). We applied Receptor-Enriched Analysis of functional Connectivity by Targets (REACT), which uses positron emission tomography (PET) maps of molecular target density to estimate, for each target, whole-brain functional connectivity (FC) with the regions where that target is most abundant. PET maps of the dopamine transporter (DAT), noradrenaline transporter (NAT), and dopamine D_1_ and D_2/3_ receptors were modelled jointly, so that each target-enriched FC map reflects that target’s unique contribution. We further compared children and adults, and explored associations with response inhibition. Methylphenidate shifted DAT- and D_2/3_-enriched FC towards less negative values and NAT-enriched FC towards less positive values, whereas D_1_-enriched FC did not change significantly. Target-enriched FC responses were broadly comparable across children and adults, and associations with response inhibition were not robust to controlling for head motion. These findings show that methylphenidate alters FC patterns linked to distinct catecholaminergic systems, supporting molecularly informed connectivity as a framework for studying the neural effects of pharmacological treatment in ADHD.

## Introduction

Methylphenidate is a widely used pharmacological treatment for attention-deficit/hyperactivity disorder (ADHD) and primarily increases catecholamine availability through blockade of the dopamine and noradrenaline transporters (DAT and NAT).^1,2^ Its therapeutic effects are thought to arise from modulation of these neural systems, but how interaction with these molecular targets translates into large-scale functional brain changes remains incompletely understood.

Resting-state functional connectivity (FC) provides a systems-level measure of coordinated brain activity and has been used to investigate the neural effects of methylphenidate in ADHD. Studies in children and adults with ADHD have reported methylphenidate-related changes in FC across distributed brain networks,^3-8^ but findings have been heterogeneous across studies, reflecting differences in participant characteristics, treatment exposure, dosing, imaging designs, and analytical approaches. ^9^ Importantly, conventional FC analyses do not directly identify which molecular targets contribute to these network-level effects.

Molecularly informed approaches can help bridge this gap by integrating neurochemical information with functional neuroimaging. Receptor-Enriched Analysis of functional Connectivity by Targets (REACT^10^) combines resting-state functional MRI with spatial maps of receptor or transporter availability to estimate functional connectivity enriched for specific molecular targets. Previous studies using REACT found that acute methylphenidate modulates DAT- and NAT-enriched FC in healthy adults,^11,12^ while studies of other pharmacological agents have demonstrated differential effects across direct and downstream molecular targets.^13^ In addition, dopamine D_1_ and D_2/3_ receptor availability has been associated with individual differences in methylphenidate-induced changes in brain function and cognition.^14,15^ Together, these findings suggest that examining both methylphenidate’s direct targets (DAT and NAT) and downstream dopamine receptors (D_1_ and D_2/3_) may provide a more comprehensive account of its effects on functional brain systems. Because these molecular distributions show varying degrees of spatial overlap, modelling them jointly may further help distinguish target-enriched contributions from effects shared across molecular systems.

Therefore, in this study we investigated the acute effects of methylphenidate on FC enriched for its direct targets (DAT, NAT) and downstream dopamine receptors (D_1_-receptor, D_2/3_-receptor) in stimulant treatment-naive children and adults with ADHD. We hypothesised that methylphenidate would elicit changes in all target-enriched FC maps, with increases in DAT-enriched FC and decreases in NAT-enriched FC based on previous work.^11,12^ Because methylphenidate improves response inhibition in ADHD, and both noradrenergic and dopaminergic signalling contribute to inhibitory control,^16-19^ we explored associations between target-enriched FC and response inhibition. In addition, because the catecholaminergic system continues to mature into adulthood and methylphenidate effects on the brain have been shown to differ between children and adults^20-23^, we explored age-group differences in methylphenidate-induced target-enriched FC changes.

## Methods

### Participants and study design

The current study includes the baseline data of the “effects of Psychotropic drugs On the Developing brain - methylphenidate” (ePOD-MPH) randomised controlled trial,^23,24^ in which 50 boys (aged 10-12 years) and 49 men (aged 23-40 years) with ADHD (all subtypes) were included. All participants were stimulant treatment-naïve. ADHD diagnosis (DSM-IV^25^) was established by an experienced psychiatrist using a structured interview in children or parents (Diagnostic Interview Schedule for Children fourth edition, DISC-IV^26^), and the Diagnostic Interview for Adult ADHD in adults (DIVA 2.0^27^). For details on recruitment and exclusion criteria, see Supplementary Materials.

The ePOD-MPH trial protocol was approved by the Central Committee on Research Involving Human Subjects (an independent registry; NL34509.000.10) and was registered at The Netherlands National Trial Register (NTR3103). All participants and parents or legal representatives of the children provided written informed consent. Here, we report on secondary analyses of the ePOD-MPH trial,^23^ investigating the acute effects of methylphenidate on FC enriched for methylphenidate’s pharmacological targets, and the relation with response inhibition. Participants underwent MR scanning before and 90 minutes after a single-dose methylphenidate-challenge (0.5 mg/kg, max. 20 mg for children, max. 40 mg for adults; Sandoz B.V., Weesp, the Netherlands). Moreover, participants performed a response inhibition task before and after the methylphenidate-challenge (see *Clinical and cognitive assessments*).

### MRI acquisition and processing

Neuroimaging data were acquired using 3T Intera or Achieva MR scanners (Philips Healthcare, Best, The Netherlands) using an 8-channel receive-only head-coil. We acquired resting-state functional MRI data using a single-shot echo-planar imaging sequence (TR/TE=2300/30 ms, voxel size=2.3×2.3×3 mm, slices=39, FA=80°, dynamics=130), and a 3D T1-weighted anatomical scan (TR/TE=9.8/4.6 ms, voxel size=0.875×0.875×1.2 mm, FOV=256×256×120, FA=8°) for registration purposes.

For details on data processing, see ^21^. In short, preprocessing was performed using fMRIPrep v1.2.3^28^ (v20.1.1 was used for the pre-challenge scan of one adult because previous runs failed) including ICA-AROMA. Next, white matter and cerebral spinal fluid (CSF) signals were regressed out and high-pass filtering (100 s) was performed using FSL. Framewise displacement (FD) was calculated from low-pass filtered motion parameter time series as described by ^29^. Volumes with FD>0.3 mm, as well as the preceding and following volume, were scrubbed from the time series. Scans were excluded from analysis if the number of volumes ≤104 after scrubbing.

### Target-enriched functional connectivity

To estimate target-enriched FC of methylphenidate’s pharmacological targets, we used high-resolution population-based positron emission tomography (PET) templates for the DAT,^30^ NAT,^31^ D_1_-receptor^32^ and D_2/3_-receptor^33^ (Figure 1; Supplementary Table S1). We selected these PET templates because DAT and NAT are the primary pharmacological targets of methylphenidate, and the availability of D_1_-receptors and D_2/3_-receptors is reported to modulate methylphenidate’s effects on the brain and cognitive functions like spatial working memory and attention.^14,15^ The PET templates were resampled to 2mm^3^ resolution and rescaled between 0 and 1. The reference regions used in the kinetic models for quantification of the PET data were masked out of the PET templates (Supplementary Table S1).

**Figure 1.**
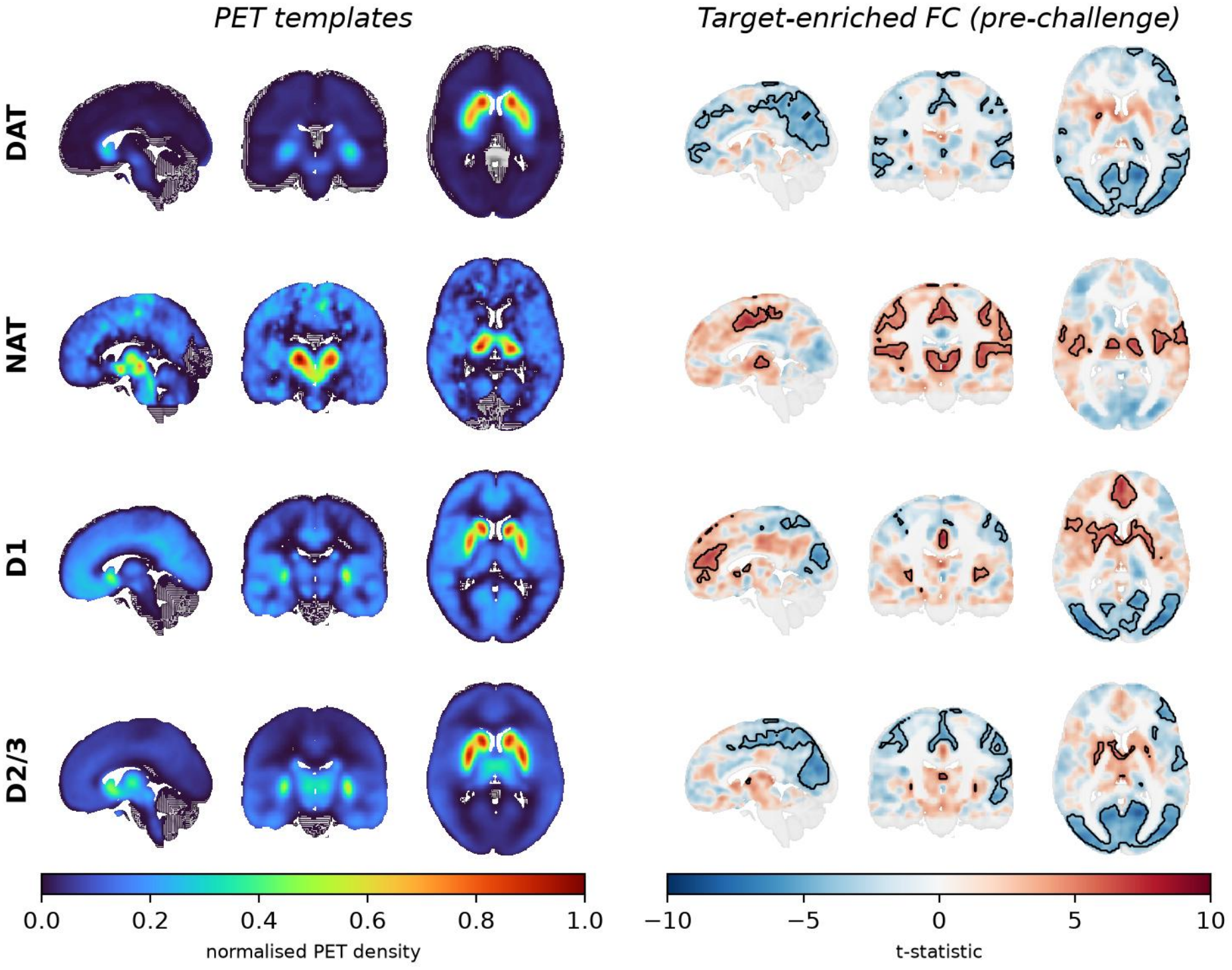
PET templates and target-enriched functional connectivity (FC) before the methylphenidate-challenge. *Top*: molecular density of the PET templates, rescaled between 0 and 1. *Bottom*: target-enriched FC maps, with motion taken into account. Significant clusters (P_FWE_<0.05) are overlaid onto the unthresholded whole-brain t-statistic map. The transparency of the unthresholded map is modulated by the t-statistic intensity, such that voxels with a lower t-statistic appear more transparent.^40,41^ Results are shown overlaid onto a 1mm MNI template brain.

Next, we applied a two-step multiple linear regression using REACT (python v3.8^10^). In the first step, the fMRI signal in the grey matter was weighted using the PET templates as spatial regressors to estimate the dominant blood-oxygenation-level-dependent (BOLD) fluctuations of the selected target-enriched functional systems at the subject level. We constructed a multiple-regression model including all PET templates in one model. In the second step, the subject-specific time series estimated in the first step were used as temporal regressors to estimate the subject-specific spatial maps of target-enriched FC before (pre-challenge) and after (post-challenge) the methylphenidate-challenge. To assess methylphenidate-induced changes in target-enriched FC, we calculated *response maps* by subtracting the pre-challenge maps from the post-challenge maps.

### Clinical and cognitive assessments

ADHD symptom severity was assessed using the Disruptive Behavior Disorder Rating-Scale (DBD-RS^34^) in children and the ADHD-Rating Scale (ADHD-RS^27^) in adults. To facilitate comparability across age groups, ADHD symptom severity sum scores were rescaled between 0 and 10 (for details, see Supplementary Methods). Intelligence quotient (IQ) was estimated using a subtest of the Wechsler Intelligence Scale for children-revised (WISC-III-R^35^) in children, and the Dutch version of the National Adult Reading Test (NART^36^) in adults.

To evaluate response inhibition, participants performed a go/nogo task (adapted from ^37^ using Pokémon cartoon characters) during MRI scanning, in addition to the resting-state functional MRI scans. Participants were instructed to press a button as fast as possible when presented with a target stimulus (go-trial), and to withhold their response to the non-target stimulus (nogo-trial). Stimulus duration was 500 ms and the interstimulus interval was 3000 ms. The task consisted of three runs with 57 trials each (43 go-trials, 14 no-go trials; nogo-trials in pseudo-randomised order), resulting in a total of 129 go-trials and 42 nogo-trials (i.e. 25% nogo-trials). We calculated dprime (i.e. the difference between the proportion of correct go-trials and the proportion of incorrect nogo-trials^38^) as a measure of performance on the response inhibition task, with higher dprime reflecting better response inhibition.

### Statistical analyses

Analysis of clinical and behavioural measures was conducted using R v4.4.1 (R Development Core Team, 2011). All voxel-wise analyses were performed using FSL Randomise v6.0.7.6 (5000 permutations; threshold-free cluster enhancement^39^; family-wise error-corrected P-values (P_FWE_)).

We conducted one-sample t-tests (positive and negative contrasts) to identify clusters with significant (methylphenidate-induced changes in) target-enriched FC for each of the investigated target maps (significance threshold P_FWE_<0.05). As target-enriched FC was estimated while taking into account the other target maps in a single multiple-regression model, we did not perform additional multiple comparison correction. Next, we conducted linear regression analyses to evaluate associations between (change in) target-enriched FC and (change in) dprime, separately for each of the target-enriched FC maps. Here, we applied a Bonferroni correction to adjust for the four target maps tested (significance threshold P_FWE_<0.0125).

In addition to the main analyses on both age groups combined (total sample), we explored potential age-related differences by running the same analyses separately per age group (children, adults). Moreover, to assess the influence of motion on our results, all analyses were run without covariates, as well as with mean FD included as covariate (or change in mean FD for the response analyses). For all analyses, we only report significant clusters encompassing at least 10 voxels. Results are visualised according to the ‘highlighting approach’ to enhance interpretation and enforce transparent reporting.^40,41^ Given recent evidence that combining spatially collinear PET templates in multivariate REACT models can inflate coefficient variance,^42^ we additionally performed a voxel-wise collinearity assessment and per-template (univariate) sensitivity analyses (Supplementary Methods).

## Results

### Participants

Of the 50 boys and 49 men that were included, 16 participants were excluded from analysis due to excessive motion (14 children), insufficient data quality (1 child), or undisclosed treatment with stimulant medication prior to study inclusion (1 adult). In addition, 2 participants were excluded from the response-analysis due to missing post-challenge data (1 child) or insufficient data quality at post-challenge (1 adult). As a result, the final sample for the REACT analysis consisted of 83 participants at pre-challenge (35 children, 48 adults), and 81 participants for the response to the methylphenidate-challenge (34 children, 47 adults).

For the participants that were included in the REACT analysis, response inhibition task (dprime) scores were missing for 20 participants (17 children, 3 adults) due to a recording error in the task. Participants with and without dprime scores did not differ in age, estimated IQ, and ADHD symptoms (children: all p>0.42; adults: all p>0.22). In addition, 2 children were excluded from the response-analysis due to missing dprime scores at post-challenge. As a result, the final sample for the associations between target-enriched FC and dprime consisted of 63 participants at pre-challenge (18 children, 45 adults), and 60 participants for the response to the methylphenidate-challenge (16 children, 44 adults).

Sample characteristics are shown in Table 1. In both age groups, performance on the response inhibition task (dprime) significantly improved after the methylphenidate-challenge (children: P=.002; adults: P<.001), and mean FD significantly decreased (children: P=.008; adults: P=.001). Furthermore, individuals with higher inattentive symptom severity performed worse on the response inhibition task at pre-challenge (total sample: R=-.27, P=.036), and individuals with higher hyperactive-impulsive symptom severity showed more methylphenidate-induced improvement on the response inhibition task (total sample: R=.27, P=.043; adults: R=.4, P=.009).

**Table 1.** Sample characteristics. Data are presented as mean ± standard deviation or median (interquartile range).

|  | **Children**<br>*n* = 35 |  |  | **Adults**<br>*n* = 48 |  |  |
|---|---|---|---|---|---|---|
| *Demographics* |  |  |  |  |  |  |
| Age (years) | 11.3 ± 0.9 |  |  | 28.5 ± 4.6 |  |  |
| IQ<sup>1</sup> | 104.1 ± 18.2 |  |  | 107.8 ± 7.5 |  |  |
| ADHD symptom severity - inattentive<sup>2</sup> | 22.2 ± 3.3 |  |  |  |  |  |
| ADHD symptom severity - Hyperactive/impulsive<sup>2</sup> | 15.5 ± 5.7 |  |  |  |  |  |
| ADHD symptom severity - Total score<sup>3</sup> |  |  |  | 32.7 ± 9.7 |  |  |
| Methylphenidate dose (mg) | 18.8 ± 2.4 |  |  | 38.3 ± 2.6 |  |  |
| *In-scanner motion & cognition* | preMPH | postMPH | statistics<sup>4</sup> | preMPH | postMPH | statistics<sup>4</sup> |
| Motion (FD, mm)<sup>5</sup> | 0.034 (0.02-0.07), *n*=35 | 0.027 (0.02-0.04), *n*=45 | t(33)=-4.61, P<.001 | 0.016 (0.01-0.02), *n*=48 | 0.013 (0.01-0.02), *n*=47 | t(46)=-3.94, P<.001 |
| dprime | 1.66±0.6, *n*=18 | 2.21±0.6, *n*=34 | t(15)=3.65, P=.002 | 2.79±0.7, *n*=45 | 3.39±0.7, *n*=45 | t(43)=7.41, P<.001 |

<sup>1</sup>Estimated using the Wechsler Intelligence Scale for children-revised (WISC-III-R) in children and the National Adult Reading Test (NART) in adults. <sup>2</sup>Disruptive Behavior Disorder-Rating Scale (DBD-RS). <sup>3</sup>ADHD-Rating Scale. <sup>4</sup>Paired-Samples T-test. <sup>5</sup>Shown for all participants that were included in the REACT analysis to evaluate (changes in) target-enriched functional connectivity. ADHD=attention-deficit/hyperactivity disorder; FD=framewise displacement; IQ=Intelligence Quotient; MPH=methylphenidate-challenge.

### Methylphenidate-induced changes in target-enriched functional connectivity

We observed significant associations between target-enriched FC and head motion (mean FD) across all investigated target maps, with participants with higher motion showing more extreme target-enriched FC values (Supplementary Figure S1; cluster information in Supplementary Table S3). For the uncorrected results from the analyses without covariates, see the Supplementary Materials (Supplementary Figure S2; cluster information in Supplementary Table S4).

When taking motion into account, before the methylphenidate-challenge we observed widespread significant positive and negative target-enriched FC for all investigated target maps (Figure 1; cluster information in Supplementary Table S5). Methylphenidate-induced changes in target-enriched FC were observed for the DAT, NAT and D_2/3_-receptor, revealing overlapping but also distinct patterns (Figure 2; cluster information in Supplementary Table S5). Specifically, DAT- and D_2/3_-enriched FC showed methylphenidate-induced increases, changing from negative to less negative target-enriched FC, whereas NAT-enriched FC showed methylphenidate-induced decreases, changing from positive to less positive target-enriched FC.

**Figure 2.**
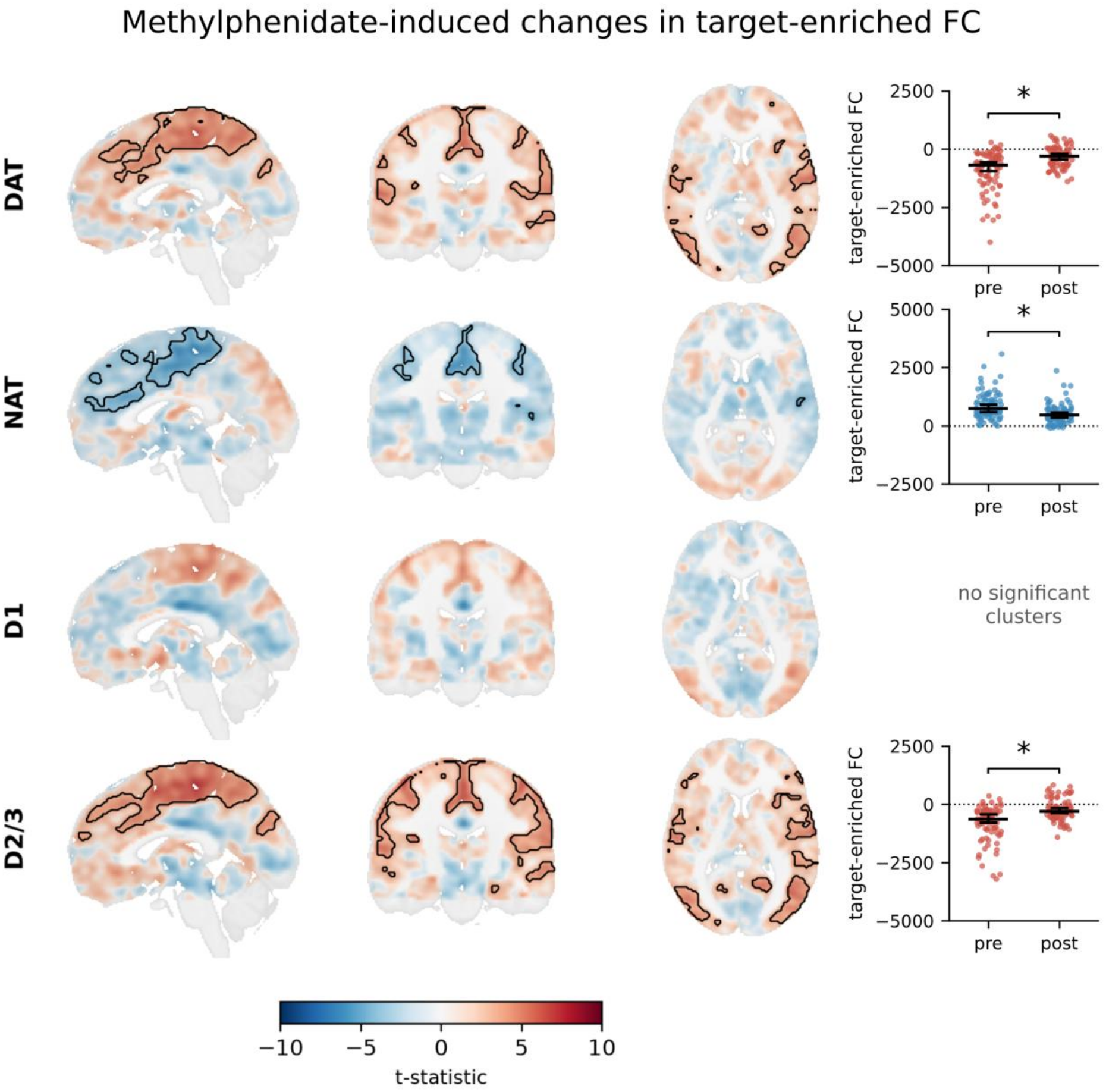
Methylphenidate-induced changes in target-enriched functional connectivity (FC). *Left*: Target-enriched FC response maps, with motion taken into account, and overlaid onto a 1mm MNI template brain. Significant clusters (P_FWE_<0.05) are overlaid onto the unthresholded whole-brain t-statistic map. The transparency of the unthresholded map is modulated by the t-statistic intensity, such that voxels with a lower t-statistic appear more transparent.^40,41^ *Right*: scatter dot plots showing the individual target-enriched FC values (mean of the significant clusters) before and after the methylphenidate-challenge. Error bars indicate median ± 95% confidence interval. *=P_FWE_<.05 for all significant clusters. pre=before methylphenidate-challenge; post=after methylphenidate-challenge.

Voxel-wise collinearity between the four PET templates was highest for D_2/3_ (VIF = 9.40) and D_1_ (VIF = 5.50), intermediate for DAT (VIF = 4.26), and lowest for NAT (VIF = 2.31) (Supplementary Table 2). Per-template sensitivity analyses yielded broadly convergent results compared with the primary multivariate model (Supplementary Figure 3). The overall pattern of DAT-, NAT-, and D_2/3_-related effects was preserved, whereas D_1_-enriched FC additionally showed a methylphenidate-induced change in the univariate analysis, suggesting sensitivity of this effect to the modelling approach.

**Figure 3.**
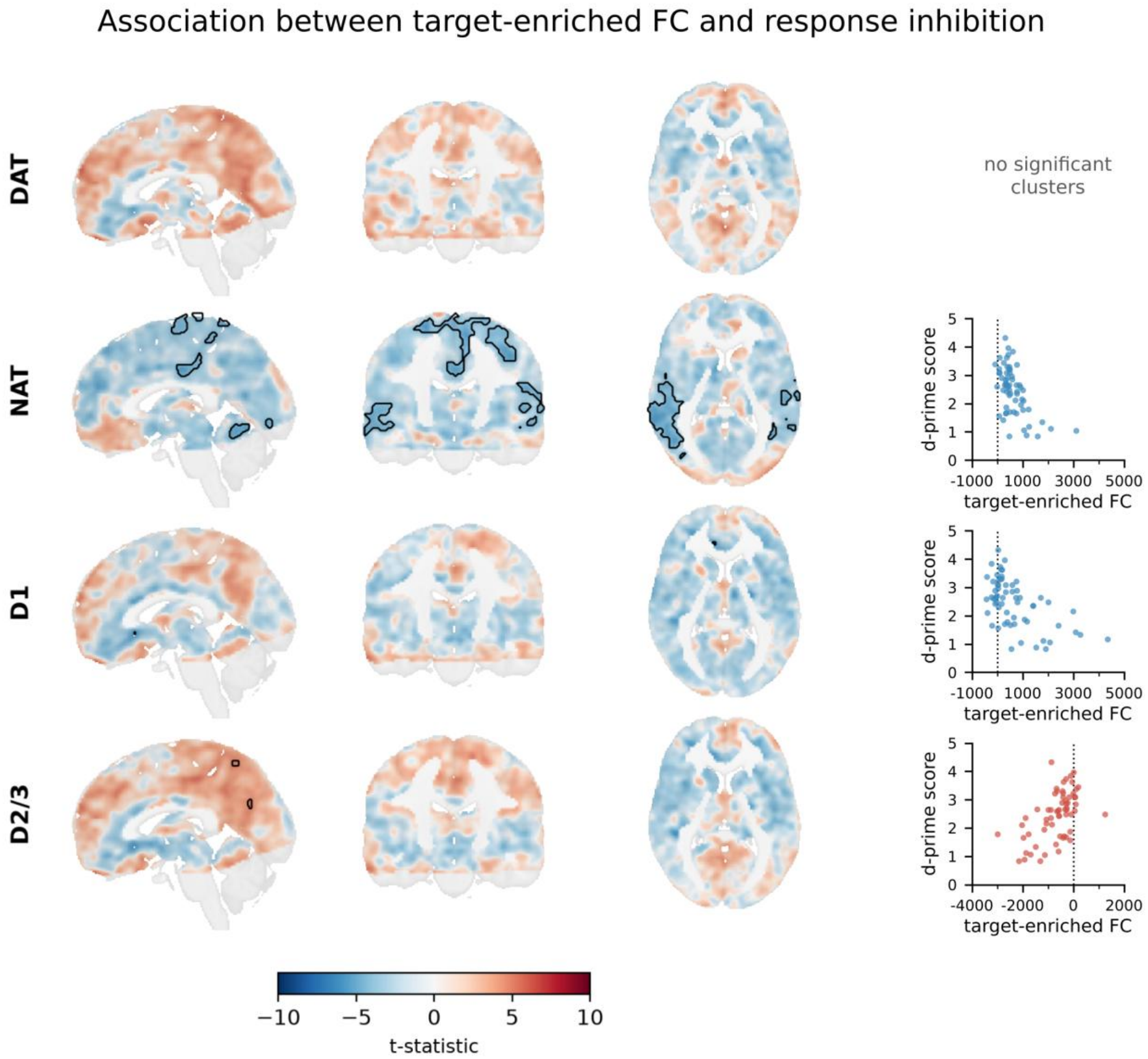
Relations between target-enriched functional connectivity (FC) and response inhibition. *Left*: target-enriched FC maps, for the models without covariates, overlaid onto a 1mm MNI template brain. Significant clusters (P_FWE_<0.0125) are overlaid onto the unthresholded whole-brain t-statistic map. The transparency of the unthresholded map is modulated by the t-statistic intensity, such that voxels with a lower t-statistic appear more transparent.^40,41^ Note that these associations were not robust when running the same models with motion as a covariate. *Right*: scatter dot plots showing the associations between target-enriched FC values (mean of the significant clusters) and dprime. The dotted vertical line indicates a target-enriched FC value of 0.

### Relations between target-enriched functional connectivity and response inhibition

Before the methylphenidate-challenge, we identified significant associations between dprime and FC enriched for the NAT, D_1_-receptor, and D_2/3_-receptor (Figure 3; cluster information in Supplementary Table S6), although these associations were not robust when taking motion into account. We found no significant associations between methylphenidate-induced changes in target-enriched FC and dprime for any of the investigated target maps.

### Age-related effects of methylphenidate

For all target maps, patterns of target-enriched FC and its associations with dprime were generally similar across age groups, although fewer and smaller significant clusters emerged in the analyses split by age group (likely related to the smaller sample size, the minimum detectable effect sizes in the stratified analyses were moderate-to-large; see Supplementary Results). For comparison of results across age groups, see Supplementary Figures S4-S11.

### 3.1 Associations of motion with clinical and cognitive measures

Since we identified associations between target-enriched FC and motion, we additionally evaluated whether motion was associated with the clinical and cognitive outcomes (Supplementary Table S7). Before the methylphenidate-challenge, we found significant associations between motion and dprime (total sample: R=-0.57, P<.0001; adults: R=-0.39, P=.007), such that participants with higher motion performed worse on the response inhibition task. Moreover, in adults methylphenidate-induced change in motion was associated with ADHD inattentive symptom severity sum score (R=.34, P=.028), such that participants with more reduction in motion displayed less severe inattentive symptoms. We found no significant associations between motion and clinical and cognitive measures in children.

## Discussion

Methylphenidate is thought to exert its therapeutic effects through dopamine and noradrenaline transporter blockade, but how this affects functional brain networks in ADHD has remained unclear. Using receptor-informed FC in stimulant treatment-naïve children and adults with ADHD, we found that a single dose of methylphenidate shifted DAT- and D_2/3_-enriched FC towards less negative values and NAT-enriched FC towards less positive values, without detectable changes in D_1_-enriched FC. These effects were preserved when each PET template was modelled separately, with differences largely confined to the spatial extent of significant clusters. Exploratory associations with response inhibition did not survive adjustment for head motion, and responses appeared broadly similar in children and adults.

The DAT-enriched FC increases converge with previous REACT studies of acute methylphenidate in adults without ADHD.^11,12^ They also align with seed-based studies across ADHD and controls reporting methylphenidate-induced changes in striatal FC with sensorimotor, frontal, temporal and occipital cortices.^4,21,43,44^ Because the DAT template is weighted towards the striatum, our findings indicate that methylphenidate attenuates the negative coupling between striatally dominated dopaminergic fluctuations and widespread cortical regions. The dopaminergic specificity of this signal is supported by the observation that DAT-enriched FC, and not NAT-enriched FC, tracks individual differences in dopamine synthesis and reward prediction error signalling.^12^ The correspondence between our ADHD sample and healthy adult samples suggests that this dopaminergic response is shared across individuals with and without ADHD. Van den Bosch & Cools^12^ additionally reported DAT-enriched FC decreases in the thalamus and other subcortical regions, which we did not replicate. Several factors may contribute to this discrepancy: our study differed in population, used weight-adjusted dosing that resulted in higher doses in most adults, applied a pre–post instead of a placebo-controlled design, and used a PET-based instead of a SPECT-based DAT template.^11^

The NAT-enriched FC decrease is consistent with substantial NAT occupancy by methylphenidate at clinically relevant doses.^1^ It contrasts with the absence of NAT-enriched FC changes after a fixed 20 mg dose in healthy adults^11^ despite the use of the same NAT template, possibly reflecting greater NAT engagement at our higher dose. Of the four templates, NAT showed the least spatial overlap with the others, and the decrease was also present when the NAT template was modelled on its own, making it unlikely that the effect arises from variance shared with dopaminergic systems. The NAT effect was centred on sensorimotor, anterior cingulate and opercular cortices, regions in which NAT-enriched FC was positive before the challenge. Although both reflected a weakening of pre-challenge coupling, the NAT effects differed from the dopaminergic effects in location and sign, suggesting that methylphenidate affects noradrenergic and dopaminergic systems in partly distinct ways.

Beyond the direct targets, methylphenidate increased D_2/3_-enriched FC in sensorimotor, lateral occipital and inferior frontal cortices and in the hippocampus and amygdala, whereas D_1_-enriched FC was unaffected. This dissociation is compatible with the higher affinity of D_2/3_-receptors for dopamine,^45-47^ which may allow a methylphenidate-induced dopamine increase to engage D_2/3_-related circuits preferentially. In line with findings by Manza et al.,^14,15^ we can speculate that regions with relatively higher D_2/3_-receptor availability (e.g. sensorimotor cortices) may be more responsive to methylphenidate than regions with relatively higher D_1_-receptor availability (e.g. association cortices). Nonetheless, a methodological account should also be considered. D_1_- and D_2/3_-templates share a pronounced striatal distribution and were the most collinear templates in our model, so variance common to both may have been attributed to D_2/3_-enriched FC, although our sensitivity analysis suggest otherwise.

Consistent with clinical expectations and literature on response inhibition in ADHD,^48,49^ we found that higher ADHD symptom severity was associated with worse pre-challenge response inhibition scores and greater methylphenidate-induced improvement in response inhibition. Before the challenge, poorer response inhibition was associated with more extreme NAT-enriched FC in sensorimotor and temporoparietal cortices, D_1_-enriched FC in the subcallosal cortex, and D_2/3_-enriched FC in superior parietal and precuneus cortices. These regions overlap with frontoparietal and fronto-basal ganglia circuits implicated in inhibitory control,^16-19^ but none of the associations survived adjustment for motion, and none were observed for methylphenidate-induced changes. In this sample, methylphenidate-induced changes in target-enriched FC therefore did not account for individual differences in the behavioural response to methylphenidate. The attenuation after motion adjustment is difficult to interpret, because motion was itself strongly associated with task performance, and adjusting for it, therefore, removes behaviourally relevant variance along with artefacts. Motion inflates short-range and attenuates long-range FC,^50^ but it also partly reflects stable traits: it relates to long-range connectivity supporting cognitive control,^51^ remains elevated in individuals with remitted ADHD,^52^ and might share genetic influences with ADHD symptoms.^53^ Separating artefact from biologically meaningful information is, therefore, a general challenge for FC studies in ADHD. The brain–behaviour associations reported here should be regarded as hypothesis-generating, particularly given the reduced sample available for these analyses because of missing task data and motion, most of which concerned children.

Earlier work in this cohort demonstrated age-dependent effects of methylphenidate on striatal dopaminergic function, cerebral perfusion and FC^21-23^. Target-enriched FC responses to an acute methylphenidate-challenge, by contrast, appeared similar across children and adults, providing no evidence for age-dependent effects. Groups were compared qualitatively and not by formal interaction tests, the stratified analyses were powered only for moderate-to-large effects, and motion-related exclusions were concentrated in children, which may have selected for less affected children. Finally, all analyses relied on PET templates from healthy adults, which may not capture developmental or ADHD-related differences in molecular architecture. Dopaminergic systems mature considerably during adolescence, especially in the prefrontal cortex,^14,54-56^ whereas noradrenergic innervation matures earlier.^57,58^

A key strength of this study is the inclusion of stimulant treatment-naive participants, as prior stimulant medication has been shown to influence DAT and D_1_-receptor expression.^59-61^ However, these findings should be interpreted in light of what the REACT method can establish. The approach identifies functional systems whose spatial organisation corresponds to the distribution of a molecular target, but it does not measure target occupancy, neurotransmitter release or receptor signalling. Our results therefore show that methylphenidate alters FC patterns aligned with catecholaminergic targets, without demonstrating that these targets mediate the changes. Another limitation is the restricted sample size for the associations with response inhibition and the stratified analysis per age group. Additionally, our sample consisted of Dutch (predominantly white) male participants within a narrow age range and lacked a typically developing control group, limiting generalisability of this study’s findings. Finally, our findings concern the acute response to a single dose and cannot be assumed to reflect the neural effects of sustained stimulant treatment.

## Conclusions

In conclusion, an acute dose of methylphenidate reduced the magnitude of FC associated with dopamine transporter, D_2/3_-receptor and noradrenaline transporter distributions in treatment-naïve individuals with ADHD. These effects were robust across modelling approaches, and the dopaminergic effects were consistent with findings in healthy adults. Molecularly informed connectivity offers opportunities to link the pharmacology of ADHD treatment to its systems-level effects on the brain. Placebo-controlled designs, age-appropriate molecular templates and individual PET measures of target engagement will be needed to establish which of these changes are specific to methylphenidate and whether they relate to clinical response.

## Data Availability

The code used is available on https://github.com/Schrantee-lab. The data used in this study are not openly available due to privacy restrictions and are available from the corresponding author on reasonable request and execution of a data use agreement.

## Acknowledgements

This project was funded by a Stichting Management Apothekers en de Gezondheidszorg (STIMAG) grant, a personal research grant awarded to LR by the Academic Medical Center, University of Amsterdam, and 11.32050.26ERA-NET PRIOMEDCHILD FP 6(EU). We thank all participants and their parents for their contribution to this study, and all students that helped with data collection and analysis.

## Author contributions

Conceptualisation: ZP, LR, HMG, LD, AS; Formal analysis and investigation: ZP, DB, AS; Funding acquisition: ZP, LR, HMG, LD, AS; Supervision: LR, HMG, LD, AS; Writing – original draft preparation: ZP, AS; Writing – review and editing: ZP, LR, HMG, DB, LD, AS.

