## Supplementary Materials for "Methylphenidate-induced changes in target-enriched functional connectivity in ADHD"

*Overview of Supplementary Materials*

**Supplementary Methods**

- Participants recruitment and exclusion criteria
- Rescaling of ADHD symptom severity sum scores
- Spatial collinearity of PET templates
- Univariate (per-map) REACT sensitivity analysis
- **Table S1**. Details about the selected PET templates

**Supplementary Results**

- Effect size calculations for stratified analyses
- **Table S2**. Voxel-wise Pearson correlation matrix between PET templates.
- **Figure S1.** Associations between motion and target-enriched FC
- **Figure S2.** Target-enriched FC without covariates (total sample)
- **Figure S3**. Full correlation pre-MPH + response with mean FD (split by map)
- **Figures S4–S11**. Age-dependent effects of methylphenidate
- **Table S3.** Cluster info: target-enriched FC vs. motion
- **Table S4.** Cluster info: REACT models without covariates
- **Table S5.** Cluster info: REACT models with motion as covariate
- **Table S6.** Cluster info: target-enriched FC vs. response inhibition (no covariates)
- **Table S7.** Spearman correlations of motion with clinical/cognitive measures

*Supplementary Methods*

**Participants recruitment and exclusion criteria**

A total of 50 boys (aged 10-12 years) and 49 men (aged 23-30 years) were included in the initial randomised controlled trial of the “effects of Psychotropic drugs On the Developing brain - methylphenidate” project (ePOD-MPH RCT). All participants were diagnosed with attention-deficit/hyperactivity disorder (ADHD; all subtypes) and eligible for pharmacological therapy (with methylphenidate). Participants were recruited through clinical programs at the Department of Child and Adolescent Psychiatry at Triversum (Alkmaar, the Netherlands), De Bascule Academic Center for Child and Adolescent Psychiatry (Amsterdam, the Netherlands), and PsyQ mental health facility (The Hague, the Netherlands).

Exclusion criteria included comorbid axis I psychiatric disorders requiring treatment with medication at study entry, a history of major neurological or medical illness or clinical treatment with drugs influencing the dopamine system (for adults before 23 years of age), such as stimulants, neuroleptics, antipsychotics, and/or $D_{2/3}$ agonists. Moreover, participants with estimated IQ < 80 were excluded (assessed using a subtest of the Wechsler Intelligence Scale for children-revised (WISC-III-R[1]) in children, and the National Adult Reading Test (NART[2]) in adults). For more details about the participant recruitment and exclusion criteria, see [3,4].

**Rescaling of ADHD symptom severity sum scores**

ADHD symptom severity was assessed using the Dutch versions of the Disruptive Behavior Disorder Rating-Scale (inattentive and hyperactive-impulsive subscales; DBD-RS[5]) in children and the ADHD-Rating Scale (ADHD-RS[6]) in adults. The inattentive and hyperactive-impulsive subscales of the DBD-RS each consist of 9 items that are scored on a 4-point likert scale (0=never or rarely, 1=sometimes, 2=often, 3=very often), yielding symptom severity sum scores ranging between 0-27. The ADHD-RS consists of 23 items (11 inattentive items, 12 hyperactive-impulsive items) that are scored on a 4-point likert scale (0=never or rarely, 1=sometimes, 2=often, 3=very often). The scores of these 23 items are combined into 18 characteristics of ADHD, yielding symptom severity sum scores ranging between 0-27.

Symptom severity sum scores were rescaled by setting the range from 0-10. Here, a score of 0 corresponds to an original score of 0, and a score of 10 corresponds to an original score of 27. For consistency across analyses, we used the rescaled symptom severity scores in the analyses with the total sample as well as the analyses stratified per age group (children, adults).

**Spatial collinearity of PET templates**

To assess spatial collinearity among the four PET receptor/transporter templates used in the REACT analysis (DAT, NAT, D1, D2/3), we computed a voxel-wise Pearson correlation matrix and variance inflation factors (VIF) between the four templates. Template values were extracted at every voxel within the grey-matter analysis mask used for the REACT analysis. VIF was calculated for each template by regressing it on the remaining three templates within the mask ($VIF = 1 / (1 - R^2)$); a VIF above 5 was considered indicative of relevant collinearity and above 10 of severe collinearity. This voxel-based approach mirrors the parcellated approach reported by [7], adapted to our voxel-based analysis pipeline. Analyses were performed in Python (nibabel, pandas, statsmodels).

**Univariate (per-map) REACT sensitivity analysis**

As a sensitivity analysis motivated by [7], who showed that combining spatially collinear PET templates in a multivariate REACT regression can inflate variance and destabilise coefficient estimates, we re-ran the REACT one-sample t-tests (pre-challenge and MPH-response target- enriched functional connectivity) separately per PET template (DAT, NAT, D1, D2/3), in addition to the primary multivariate (maps-combined) model, for the combined age groups. All univariate models included mean framewise displacement (meanFD) as a covariate, consistent with the primary analyses reported in the main text. Group-level statistics were conducted in the same way as in the main manuscript, and because the maps were compared separately, they were Bonferroni-corrected (P<0.125 considered significant).

**Table S1. Details about the selected PET templates.**

| Map | Source | Tracer | Resolution | Reference region | Sample size (*n* female) | Sample age (mean ± SD) |
|---|---|---|---|---|---|---|
| Dopamine transporter | Sasaki et al.[8] | fepe2i | 1mm | Cerebellum | 6 (0) | 31.06 ± 7.7 |
| Noradrenaline transporter | Hesse et al.[9] | MRB | 3mm | Occipital cortex* | 10 (4) | 33.3 ± 10 |
| $D_1$-receptor | Kaller et al.[10] | sch23390 | 3mm | Cerebellum | 13 (7) | 33 ± 13 |
| $D_{2/3}$-receptor | Sandiego et al.[11] | flb457 | 1mm | Cerebellum | 55 (35) | 32.45 ± 9.69 |

*We used Brainnetome Atlas parcellation numbers 189, 190, 203 and 204 as reference regions.

### *Supplementary Results*

**Effect size calculations for stratified analyses**

Calculations were performed using G*Power (v.3.1.9.6.) for the analyses without covariates.

*REACT analyses*: based on 80% statistical power, alpha=0.05 and sample size n=34 (children) and n=47 (adults), the two-tailed one-sample t-tests were able to detect moderate Cohen's *d* effect sizes of at least 0.50 for children and 0.42 for adults.

*Associations with dprime*: based on 80% statistical power, alpha=0.0125 and sample size n=16 (children) and n=44 (adults), the linear regression models were able to detect moderate-to-large $f^2$ effect sizes of at least 0.89 for children and 0.27 for adults. These effect sizes correspond to a minimum detectable association coefficient r of 0.66 for children and 0.26 for adults.

**Spatial collinearity between PET templates**

Voxel-wise Pearson correlations between the four PET receptor/transporter templates, computed within the grey-matter analysis mask, are shown in **Supplementary Table S2**. Correlations were highest between DAT and D2/3 (r = .85), D1 and D2/3 (r = .81), and DAT and D1 (r = .71), whereas NAT showed comparatively low correlations with the other three templates (r = .08–.26).

Variance inflation factors (VIF) confirmed this pattern: VIF was highest for $D_{2/3}$ (VIF = 9.40), followed by $D_1$ (VIF = 5.50) and DAT (VIF = 4.26), while NAT showed the lowest VIF (VIF = 2.31). Using conventional thresholds (VIF > 5 indicating relevant, and VIF > 10 severe, collinearity), D2/3 and D1 showed the most relevant collinearity with the other templates, DAT was intermediate, and NAT was essentially uncorrelated with the other templates; consistent with the pattern reported by [7].

**Supplementary Table S2. Voxel-wise Pearson correlation matrix between PET templates.**

| | **DAT** | **NAT** | **$D_1$** | **$D_{2/3}$** |
|---|---|---|---|---|
| **DAT** | 1.00 | .08 | .71 | .85 |
| **NAT** | .08 | 1.00 | .25 | .26 |
| **$D_1$** | .71 | .25 | 1.00 | .81 |
| **$D_{2/3}$** | .85 | .26 | .81 | 1.00 |

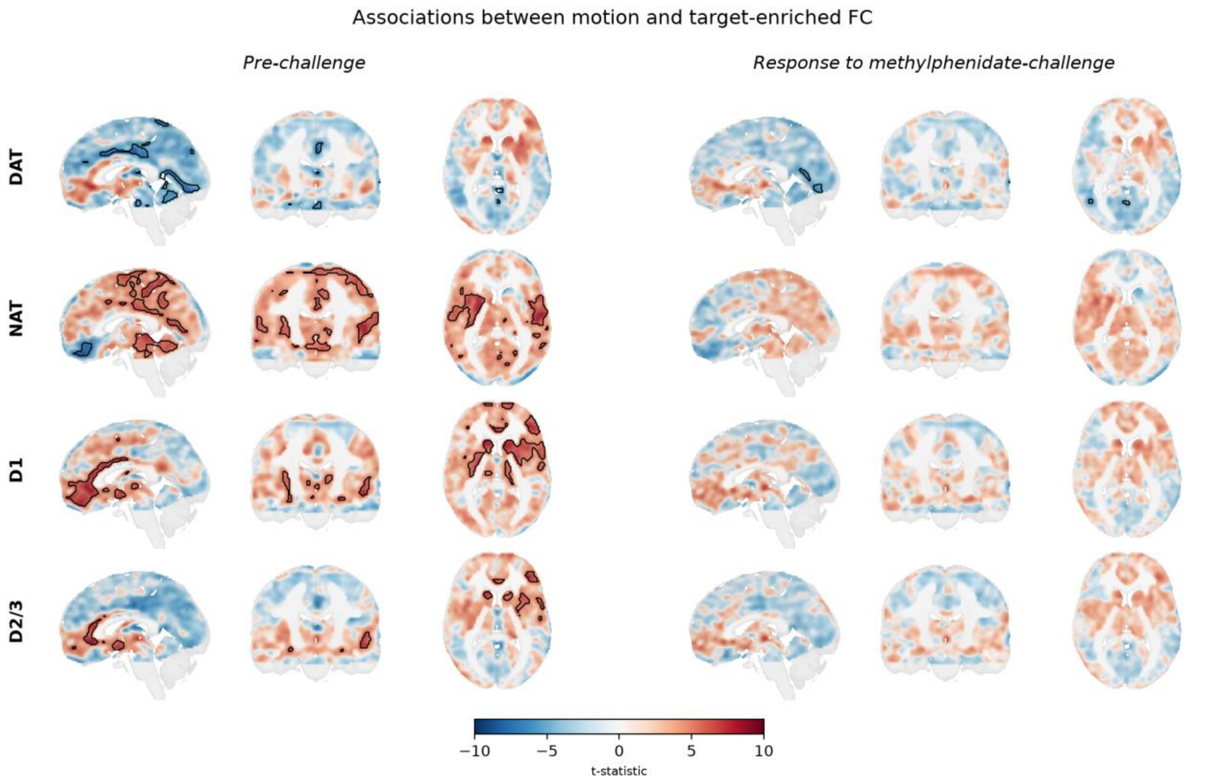


**Figure S1. Associations between motion and target-enriched FC identified using the one-sample t-tests with mean framewise displacement (FD) included as a covariate.** Target-enriched FC maps overlaid onto a 1mm MNI template brain. Significant clusters ($P_{FWE}$<0.05) are overlaid onto the unthresholded whole-brain t-statistic map. The transparency of the unthresholded map is modulated by the voxel intensity, such that voxels with a lower t-statistic appear more transparent.[12,13]

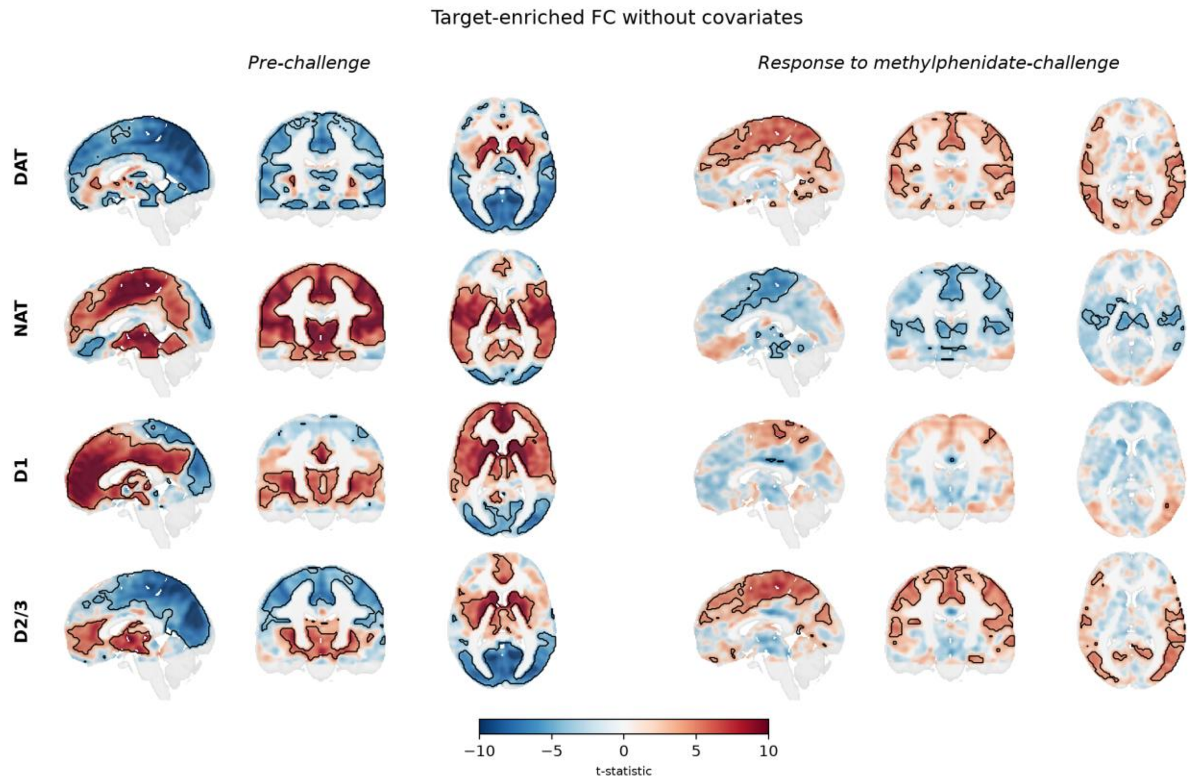


**Figure S2. Target-enriched functional connectivity (FC) without covariates, for the total sample.** *Left:* Before the methylphenidate-challenge. *Right*: Methylphenidate-induced changes. Significant clusters ($P_{FWE}$<0.05) are overlaid onto the unthresholded whole-brain t-statistic map. The transparency of the unthresholded map is modulated by the voxel intensity, such that voxels with a lower t-statistic appear more transparent.[12,13] Results are shown overlaid onto a 1mm MNI template brain. Scatter dot plots showing the individual target-enriched FC values (mean of the significant clusters) before and after the methylphenidate-challenge. Error bars indicate median ± 95% confidence interval. *=$P_{FWE}$<.05 for all significant clusters.
.

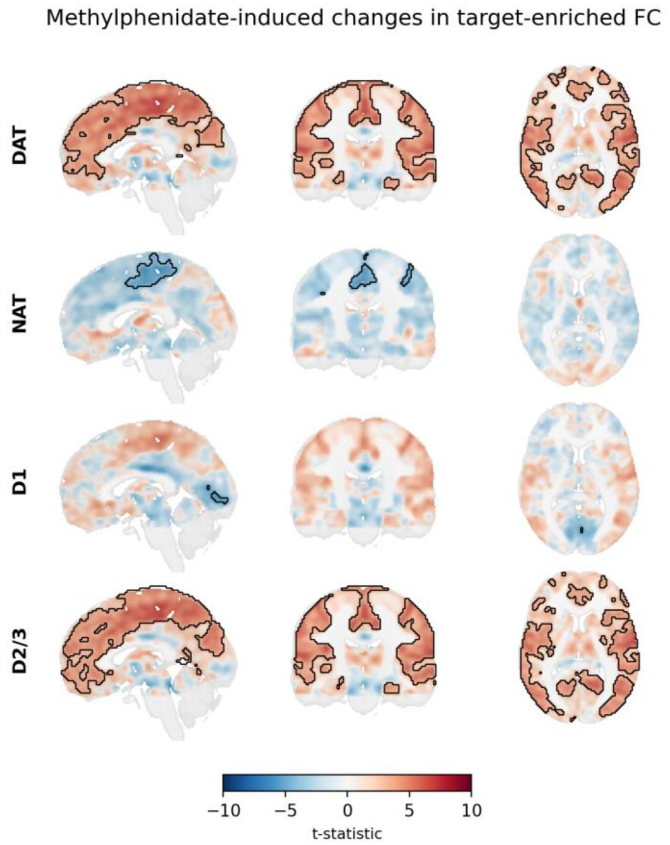


**Figure S3. Methylphenidate-induced changes in target-enriched functional connectivity (FC) per map tested separately.** Target-enriched FC response maps, with motion taken into account, and overlaid onto a 1mm MNI template brain. Significant clusters ($P_{FWE}$<0.05) are overlaid onto the unthresholded whole-brain t-statistic map.

### Age-dependent effects of methylphenidate on target-enriched functional connectivity and relations with response inhibition

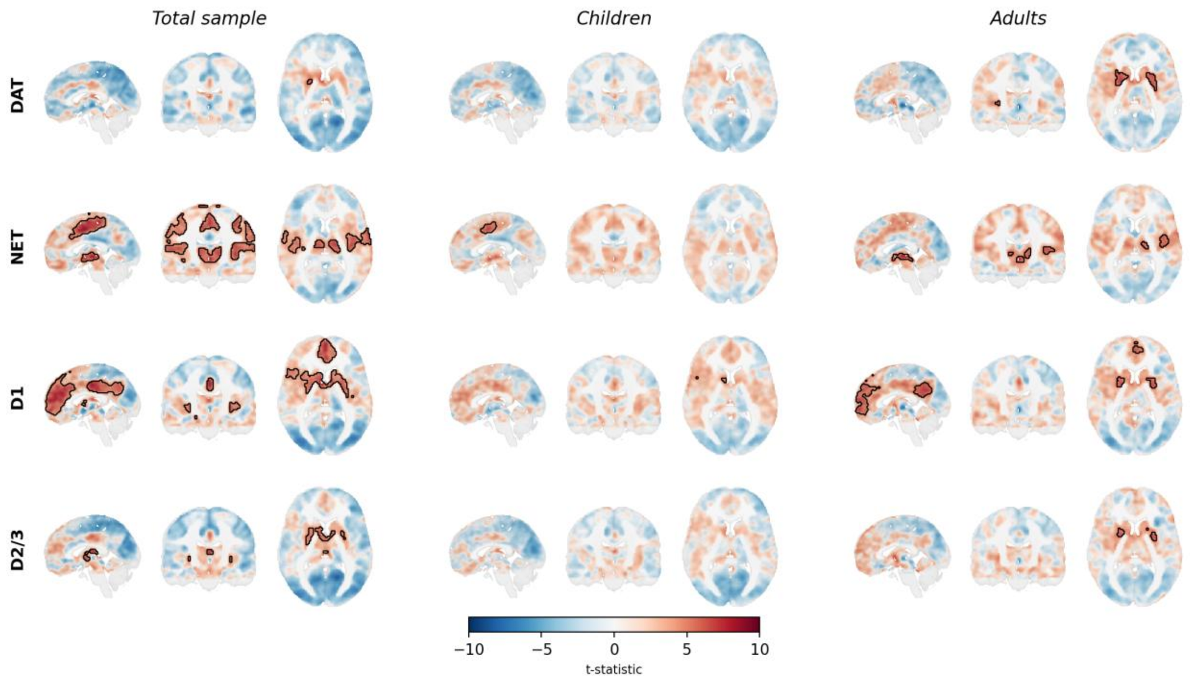


**Figure S4. Comparison of pre-challenge target-enriched functional connectivity (FC) across age groups (total sample, children, adults).** Results are shown overlaid onto a 1mm MNI template brain, for the analyses with motion (mean framewise displacement) included as covariate. Significant clusters ($P_{FWE}$<0.05) are overlaid onto the unthresholded whole-brain t-statistic map. The transparency of the unthresholded map is modulated by the voxel intensity, such that voxels with a lower t-statistic appear more transparent.[12,13]

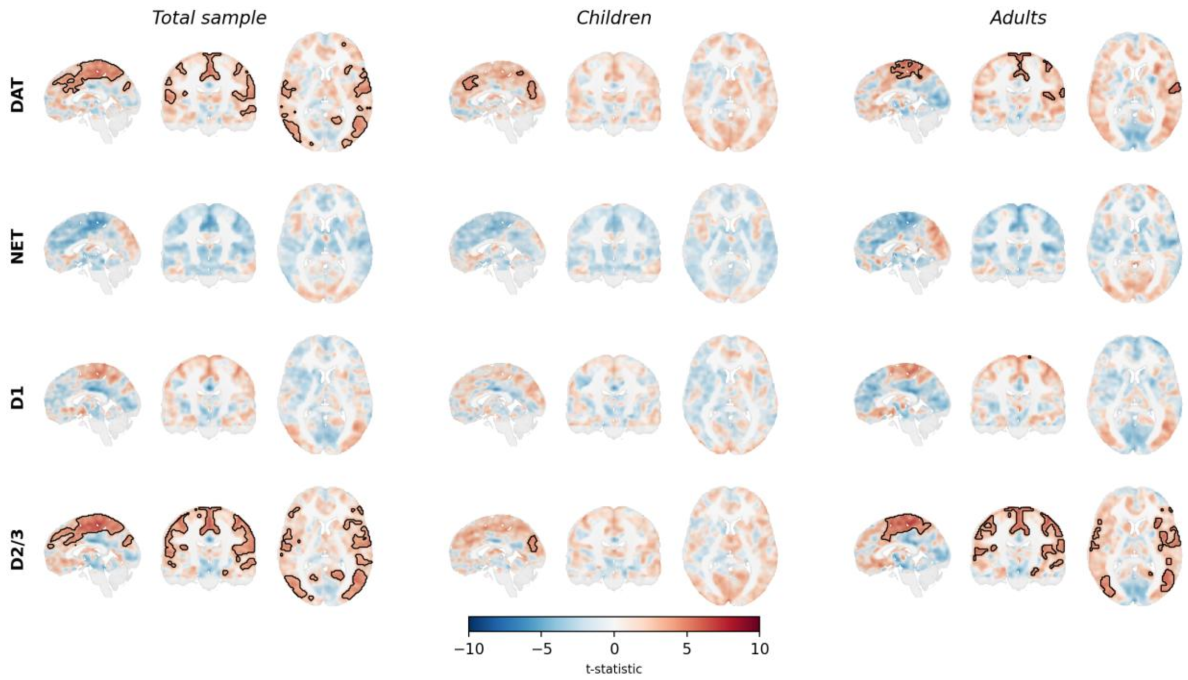


**Figure S5. Comparison of methylphenidate-induced changes in target-enriched functional connectivity (FC) across age groups (total sample, children, adults).** Results are shown overlaid onto a 1mm MNI template brain, for the analyses with motion (mean framewise displacement) included as covariate. Significant clusters ($P_{FWE}$<0.05) are overlaid onto the unthresholded whole-brain t-statistic map. The transparency of the unthresholded map is modulated by the voxel intensity, such that voxels with a lower t-statistic appear more transparent.[12,13]

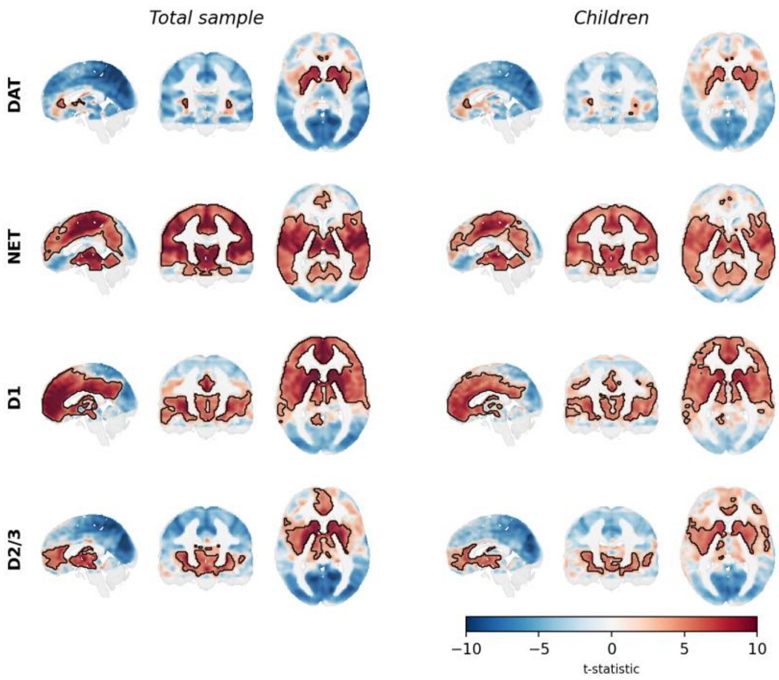


**Figure S6. Comparison of pre-challenge target-enriched functional connectivity (FC) across age groups (total sample, children, adults).** Results are shown overlaid onto a 1mm MNI template brain, for the analyses *without covariates*. Significant clusters ($P_{FWE}$<0.05) are overlaid onto the unthresholded whole-brain t-statistic map.

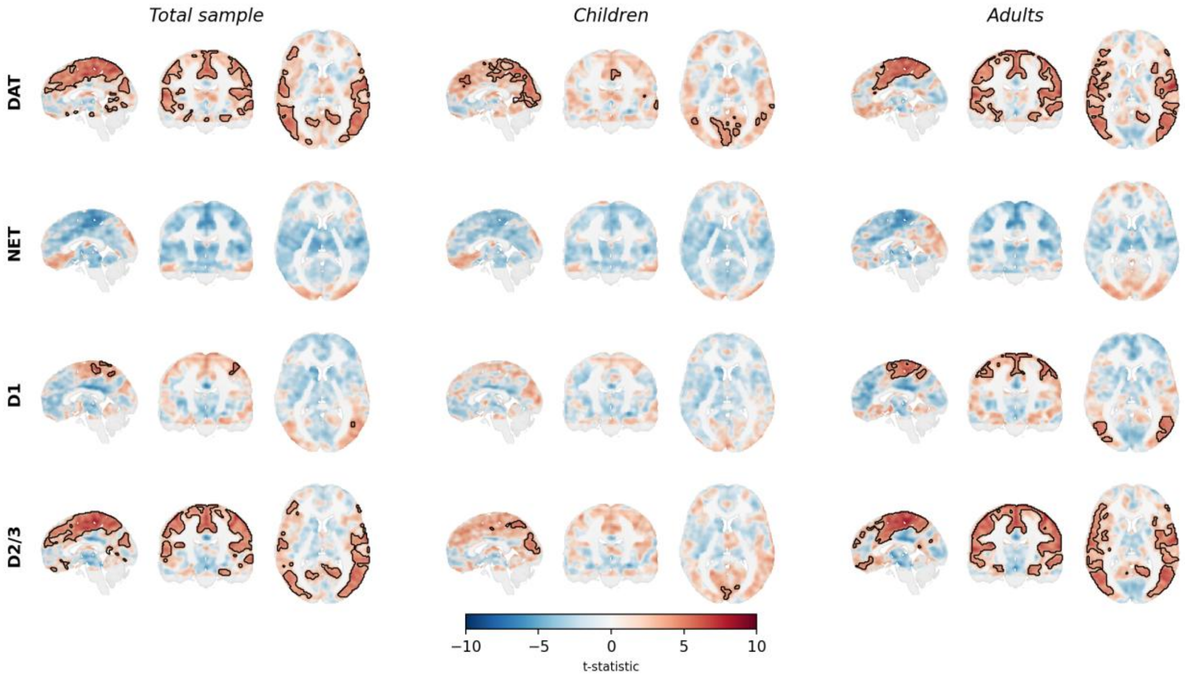


**Figure S7. Comparison of methylphenidate-induced changes in target-enriched functional connectivity (FC) across age groups (total sample, children, adults).** Results are shown overlaid onto a 1mm MNI template brain, for the analyses *without covariates*. Significant clusters ($P_{FWE}$<0.05) are overlaid onto the unthresholded whole-brain t-statistic map.

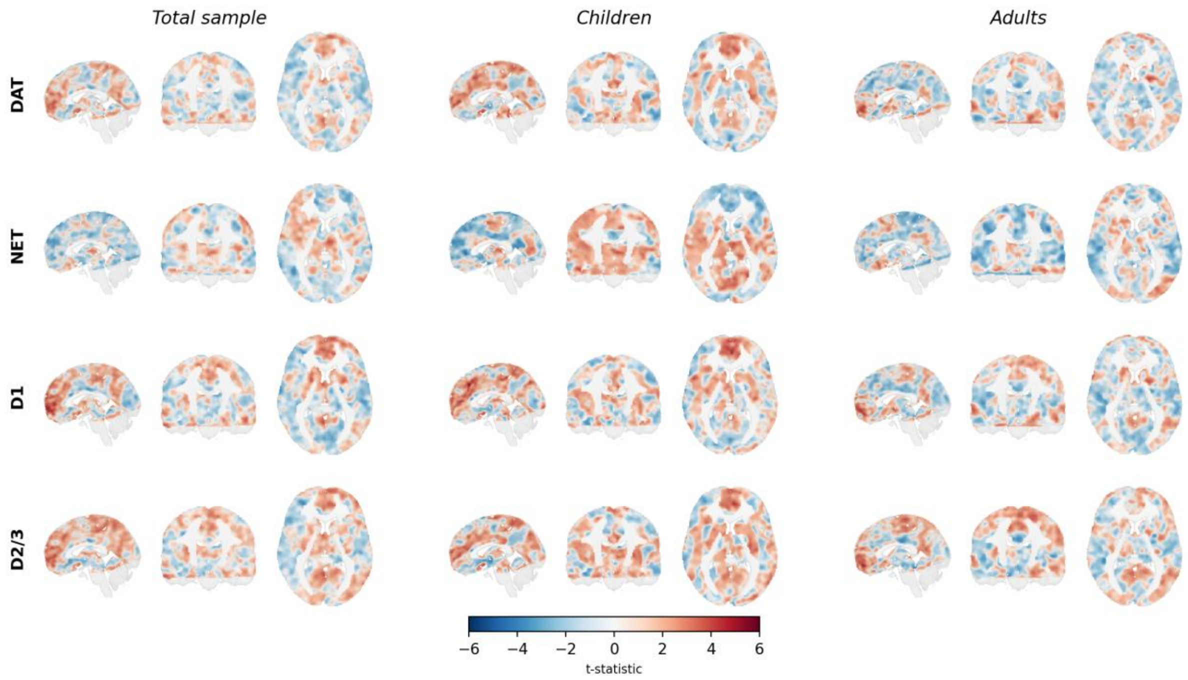


**Figure S8. Comparison of associations between pre-challenge target-enriched functional connectivity (FC) and response inhibition across age groups (total sample, children, adults).** Results are shown overlaid onto a 1mm MNI template brain, for the analyses with motion (mean framewise displacement) included as covariate. No significant clusters ($P_{FWE}$<0.05) were identified, so only the unthresholded whole-brain t-statistic map is shown.

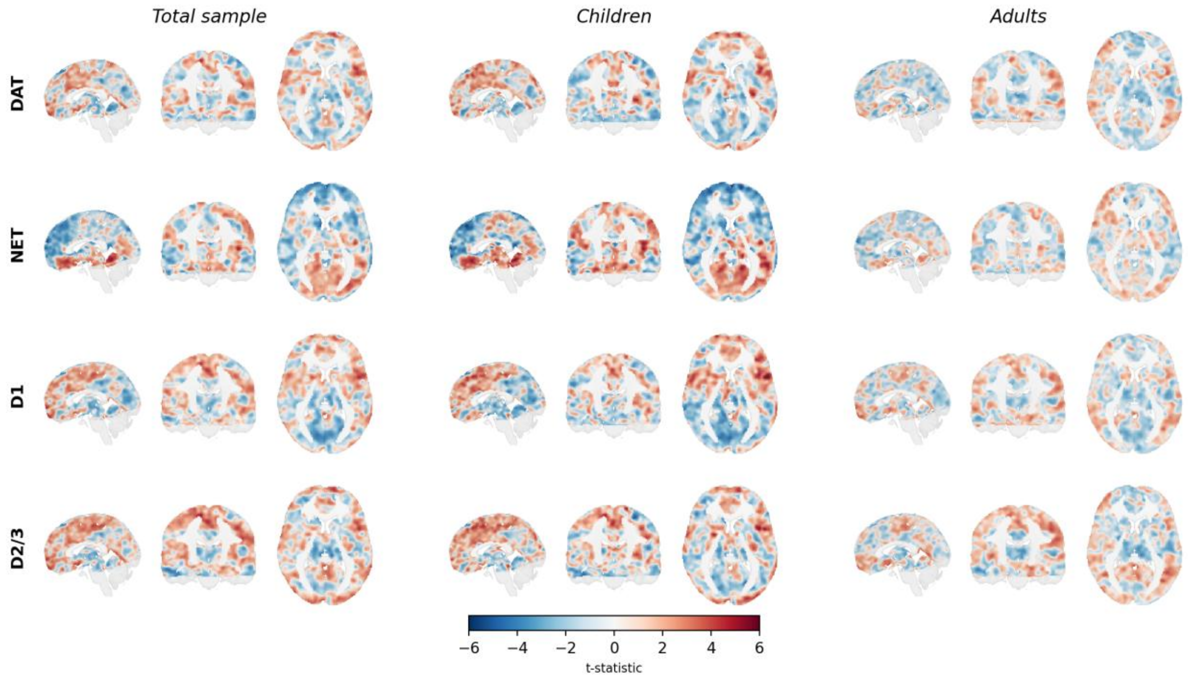


**Figure S9. Comparison of associations between methylphenidate-induced changes in target-enriched functional connectivity (FC) and response inhibition across age groups (total sample, children, adults).** Results are shown overlaid onto a 1mm MNI template brain, for the analyses with motion (mean framewise displacement) included as covariate. No significant clusters ($P_{FWE}$<0.05) were identified, so only the unthresholded whole-brain t-statistic map is shown.

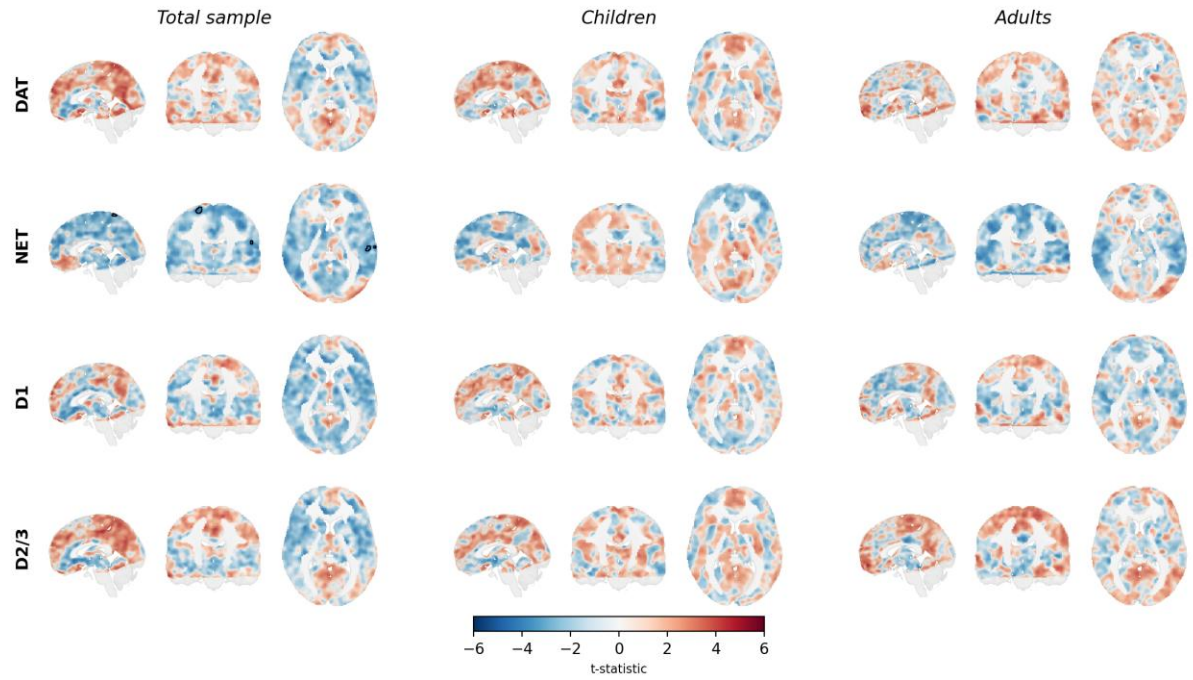


**Figure S10. Comparison of associations between pre-challenge target-enriched functional connectivity (FC) and response inhibition across age groups (total sample, children, adults).** Results are shown overlaid onto a 1mm MNI template brain, for the analyses *without covariates*. Significant clusters ($P_{FWE}$<0.05) are overlaid onto the unthresholded whole-brain t-statistic map.

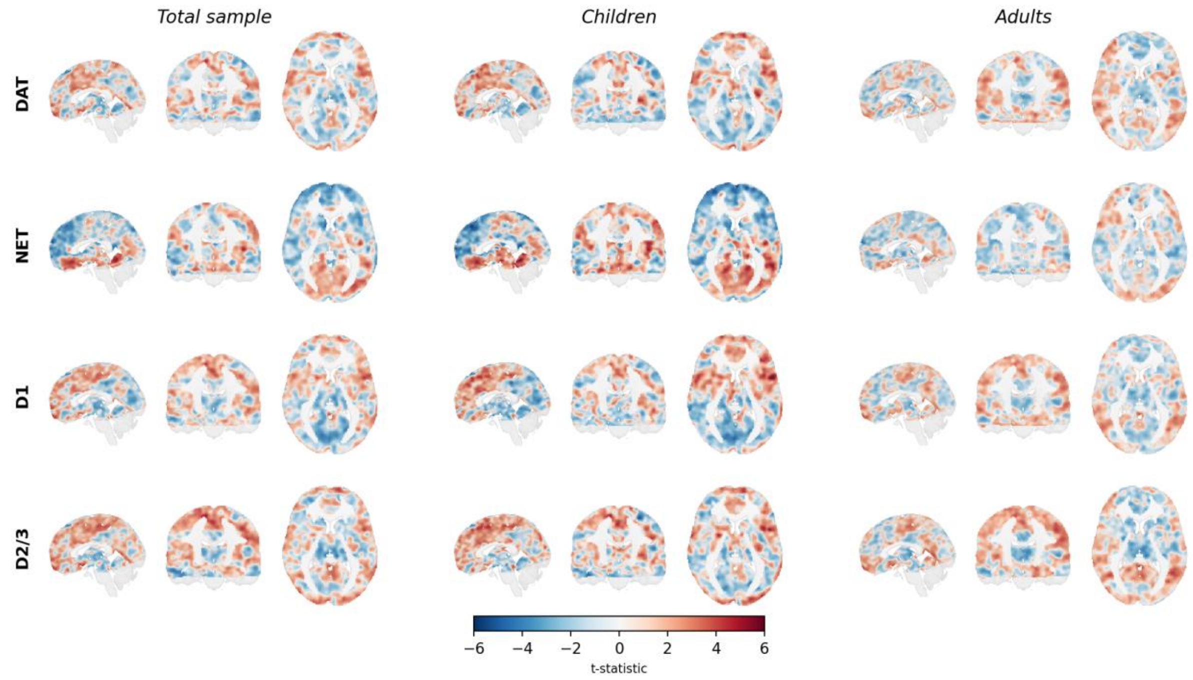


**Figure S10. Comparison of associations between methylphenidate-induced changes in target-enriched functional connectivity (FC) and response inhibition across age groups (total sample, children, adults).** Results are shown overlaid onto a 1mm MNI template brain, for the analyses *without covariates*. No significant clusters ($P_{FWE}$<0.05) were identified, so only the unthresholded whole-brain t-statistic map is shown.

**Table S2. Cluster information showing significant associations between target-enriched FC and motion.** The table shows significant positive/negative associations of framewise displacement (FD) with target-enriched FC (pre-challenge) and methylphenidate-induced target-enriched FC changes (response to methylphenidate-challenge). Cluster information was extracted using FSL's autoaq. Clusters of ≥10 voxels and ≥5% overlap are reported. DAT=dopamine transporter; FC=functional connectivity; NAT=noradrenaline transporter.

| **Pre-challenge** | | | | | |
|---|---|---|---|---|---|
| | **Cluster** | **# voxels** | **Max p-value** | **Overlap (%)** | **Region** |
| *DAT-enriched FC* | | | | | |
| Negative | 16 | 18 | 0.016 | 15.9 | Frontal Orbital Cortex |
| | | | | 11.5 | Temporal Pole |
| | | | | 10.8 | Frontal Operculum Cortex |
| Negative | 16 | 18 | 0.016 | 6.4 | Inferior Frontal Gyrus |
| | | | | 25.1 | Intracalcarine Cortex |
| | | | | 13.9 | Lingual Gyrus |
| | | | | 5.0 | Occipital Pole |
| Negative | 18 | 28 | 0.041 | 60.6 | Frontal Orbital Cortex |
| | | | | 15.5 | Subcallosal Cortex |
| Negative | 19 | 34 | 0.03 | 37.5 | Cuneal Cortex |
| | | | | 32.8 | Precuneus Cortex |
| Negative | 20 | 45 | 0.024 | 92.3 | Brain-Stem |
| Negative | 21 | 195 | 0.01 | 18.6 | Superior Parietal Lobule |
| | | | | 17.7 | Postcentral Gyrus |
| | | | | 15.1 | Precuneus Cortex |
| Negative | 22 | 231 | 0.006 | 45.8 | Posterior Cingulate Gyrus |
| | | | | 34.2 | Anterior Cingulate Gyrus |
| Negative | 23 | 379 | 0.002 | 28.3 | Frontal Orbital Cortex |
| | | | | 16.8 | Subcallosal Cortex |
| | | | | 11.7 | Frontal Pole |
| Negative | 24 | 3853 | <0.001 | 11.7 | Lingual Gyrus |
| | | | | 5.7 | Inferior Temporal Gyrus |
| Negative | 24 | 3853 | <0.001 | 5.1 | Temporal Occipital Fusiform Cortex |
| *NAT-enriched FC* | | | | | |
| Positive | 15 | 12 | 0.039 | 58.3 | Inferior Lateral Occipital Cortex |
| | | | | 18.4 | Middle Temporal Gyrus |
| Positive | 16 | 12 | 0.032 | 51.6 | Superior Lateral Occipital Cortex |
| | | | | 7.8 | Superior Parietal Lobule |
| Positive | 17 | 15 | 0.023 | 83.5 | Anterior Cingulate Gyrus |
| Positive | 19 | 18 | 0.04 | 44.4 | Inferior Lateral Occipital Cortex |
| Positive | 20 | 19 | 0.025 | 57.9 | Anterior Cingulate Gyrus |
| | | | | 23.2 | Paracingulate Gyrus |
| Positive | 21 | 31 | 0.035 | 60.2 | Superior Lateral Occipital Cortex |
| Positive | 22 | 45 | 0.032 | 35.5 | Postcentral Gyrus |
| | | | | 23.8 | Precentral Gyrus |
| Positive | 23 | 48 | 0.042 | 22.7 | Inferior Temporal Gyrus |
| | | | | 21.9 | Temporal Occipital Fusiform Cortex |
| | | | | 10.3 | Posterior Temporal Fusiform Cortex |
| | | | | 8.4 | Posterior Inferior Temporal Gyrus |
| Positive | 24 | 65 | 0.038 | 28.6 | Intracalcarine Cortex |
| | | | | 9.8 | Lingual Gyrus |
| Positive | 25 | 77 | 0.038 | 22.0 | Precuneus Cortex |
| | | | | 16.0 | Supracalcarine Cortex |
| | | | | 10.2 | Intracalcarine Cortex |
| Positive | 26 | 95 | 0.022 | 26.7 | Angular Gyrus |
| | | | | 17.8 | Superior Lateral Occipital Cortex |
| | | | | 10.7 | Middle Temporal Gyrus |
| | | | | 8.3 | Inferior Lateral Occipital Cortex |
| Positive | 27 | 97 | 0.033 | 29.0 | Lingual Gyrus |
| | | | | 26.4 | Precuneus Cortex |
| | | | | 17.5 | Intracalcarine Cortex |
| Positive | 28 | 183 | 0.024 | 42.7 | Superior Lateral Occipital Cortex |
| | | | | 5.3 | Superior Parietal Lobule |
| Positive | 29 | 1155 | 0.015 | 36.6 | Superior Lateral Occipital Cortex |

| | Cluster | # voxels | Max p-value | Overlap (%) | Region |
|---|---|---|---|---|---|
| | | | | 18.5 | Angular Gyrus |
| Positive | 30 | 5172 | 0.001 | 9.1 | Right Putamen |
| | | | | 6.9 | Brain-Stem |
| | | | | 5.1 | Insular Cortex |
| Positive | 31 | 7991 | 0.001 | 11.0 | Precentral Gyrus |
| | | | | 9.9 | Postcentral Gyrus |
| Negative | 4 | 34 | 0.013 | 63.4 | Frontal Pole |
| Negative | 5 | 617 | 0.003 | 33.9 | Frontal Medial Cortex |
| | | | | 26.1 | Frontal Pole |
| | | | | 7.5 | Paracingulate Gyrus |
| *$D_1$-enriched FC* | | | | | |
| Positive | 4 | 16 | 0.047 | 63.9 | Juxtapositional Lobule Cortex |
| | | | | 5.4 | Paracingulate Gyrus |
| Positive | 5 | 11700 | <0.001 | 9.6 | Frontal Pole |
| | | | | 5.9 | Insular Cortex |
| *$D_{2/3}$-enriched FC* | | | | | |
| Positive | 6 | 42 | 0.034 | 39.5 | Frontal Pole |
| | | | | 31.0 | Frontal Medial Cortex |
| Positive | 7 | 98 | 0.026 | 70.9 | Left Caudate |
| Positive | 8 | 1364 | 0.003 | 18.4 | Anterior Cingulate Gyrus |
| | | | | 10.1 | Right Caudate |
| | | | | 6.0 | Right Putamen |
| Positive | 9 | 1601 | 0.002 | 14.4 | Left Putamen |
| | | | | 9.0 | Frontal Pole |
| | | | | 7.0 | Frontal Orbital Cortex |
| | | | | 5.9 | Insular Cortex |

**Response to methylphenidate-challenge**

| | Cluster | # voxels | Max p-value | Overlap (%) | Region |
|---|---|---|---|---|---|
| *DAT-enriched FC* | | | | | |
| Negative | 8 | 19 | 0.039 | 67.0 | Left Thalamus |
| | | | | 8.4 | Left Hippocampus |
| Negative | 9 | 24 | 0.042 | 74.6 | Inferior Lateral Occipital Cortex |
| Negative | 10 | 30 | 0.031 | 40.0 | Inferior Lateral Occipital Cortex |
| | | | | 19.1 | Inferior Temporal Gyrus |
| | | | | 14.8 | Temporal Occipital Fusiform Cortex |
| Negative | 11 | 54 | 0.035 | 26.2 | Middle Temporal Gyrus |
| | | | | 17.0 | Inferior Lateral Occipital Cortex |
| Negative | 12 | 62 | 0.026 | 29.3 | Posterior Cingulate Gyrus |
| | | | | 24.1 | Lingual Gyrus |
| Negative | 13 | 146 | 0.012 | 33.5 | Lingual Gyrus |
| | | | | 12.5 | Intracalcarine Cortex |
| | | | | 6.8 | Precuneus Cortex |
| Negative | 14 | 211 | 0.013 | 28.8 | Lingual Gyrus |
| | | | | 16.6 | Occipital Fusiform Gyrus |
| | | | | 15.9 | Temporal Occipital Fusiform Cortex |
| Negative | 15 | 408 | 0.007 | 44.5 | Lingual Gyrus |
| | | | | 14.2 | Occipital Fusiform Gyrus |

**Table S3. Cluster information for the REACT models without covariates.** The table shows significant positive/negative target-enriched FC (pre-challenge) and significant increases/decreases in target-enriched FC (response to methylphenidate-challenge). Cluster information was extracted using FSL's autoaq. Clusters of ≥10 voxels and ≥5% overlap are reported. DAT=dopamine transporter; FC=functional connectivity; NAT=noradrenaline transporter.

| **Pre-challenge** | | | | | |
|---|---|---|---|---|---|
| | **Cluster** | **# voxels** | **Max p-value** | **Overlap (%)** | **Region** |
| *DAT-enriched FC* | | | | | |
| Positive | 2 | 5054 | <0.001 | 13.5 | Right Putamen |
| | | | | 12.9 | Left Putamen |
| | | | | 6.9 | Left Caudate |
| | | | | 6.8 | Right Caudate |
| | | | | 6.2 | Insular Cortex |
| Negative | 5 | 75 | 0.019 | 34.4 | Frontal Pole |
| | | | | 20.9 | Frontal Medial Cortex |
| Negative | 6 | 87316 | <0.001 | 6.0 | Superior Lateral Occipital Cortex |
| *NAT-enriched FC* | | | | | |
| Positive | 2 | 12 | 0.044 | 34.5 | Middle Frontal Gyrus |
| Positive | 3 | 67847 | <0.001 | 5.7 | Precentral Gyrus |
| | | | | 5.0 | Postcentral Gyrus |
| Negative | 7 | 15 | 0.009 | 16.8 | Subcallosal Cortex |
| | | | | 8.5 | Right Caudate |
| | | | | 7.4 | Left Caudate |
| Negative | 8 | 2385 | <0.001 | 38.8 | Occipital Pole |
| | | | | 14.2 | Superior Lateral Occipital Cortex |
| | | | | 7.0 | Inferior Lateral Occipital Cortex |
| Negative | 9 | 2412 | 0.001 | 34.8 | Frontal Pole |
| | | | | 13.6 | Frontal Medial Cortex |
| | | | | 10.6 | Frontal Orbital Cortex |
| *$D_1$-enriched FC* | | | | | |
| Positive | 3 | 12 | 0.01 | 41.9 | Left Hippocampus |
| Positive | 4 | 21 | 0.001 | 42.4 | Superior Frontal Gyrus |
| | | | | 9.5 | Juxtapositional Lobule Cortex |
| Positive | 5 | 46189 | <0.001 | 11.3 | Frontal Pole |
| Negative | 4 | 10 | 0.019 | 41.6 | Brain-Stem |
| | | | | 55.1 | Posterior Temporal Fusiform Cortex |
| | | | | 7.5 | Posterior Parahippocampal Gyrus |
| | | | | 7.1 | Anterior Parahippocampal Gyrus |
| Negative | 6 | 26550 | <0.001 | 12.0 | Superior Lateral Occipital Cortex |
| | | | | 6.6 | Inferior Lateral Occipital Cortex |
| | | | | 5.1 | Occipital Pole |
| *$D_{2/3}$-enriched FC* | | | | | |
| Positive | 2 | 15578 | <0.001 | 6.6 | Insular Cortex |
| Negative | 12 | 11 | 0.027 | 74.3 | Frontal Pole |
| Negative | 14 | 24 | 0.04 | 84.9 | Frontal Pole |
| Negative | 15 | 30 | 0.027 | 29.7 | Middle Temporal Gyrus |
| | | | | 13.2 | Posterior Middle Temporal Gyrus |
| Negative | 16 | 639 | 0.016 | 30.9 | Frontal Orbital Cortex |
| | | | | 17.7 | Frontal Pole |
| | | | | 9.0 | Subcallosal Cortex |
| Negative | 17 | 64955 | <0.001 | 7.4 | Superior Lateral Occipital Cortex |
| | | | | 5.5 | Precentral Gyrus |
| | | | | 5.2 | Precuneus Cortex |
| **Response to methylphenidate-challenge** | | | | | |
| | **Cluster** | **# voxels** | **Max p-value** | **Overlap (%)** | **Region** |
| *DAT-enriched FC* | | | | | |
| Increase | 8 | 17 | 0.026 | 98.5 | Brain-Stem |
| Increase | 9 | 27 | 0.041 | 10.7 | Insular Cortex |
| | | | | 10.7 | Planum Polare |
| Increase | 10 | 70 | 0.036 | 19.7 | Right Amygdala |
| | | | | 12.8 | Temporal Pole |
| | | | | 8.5 | Planum Polare |

| | | | | | |
|---|---|---|---|---|---|
| | | | | 7.5 | Insular Cortex |
| | | | | 5.7 | Anterior Parahippocampal Gyrus |
| Increase | 11 | 43494 | <0.001 | 5.5 | Superior Lateral Occipital Cortex |
| *NAT-enriched FC* | | | | | |
| Decrease | 8 | 17 | 0.04 | 53.8 | Middle Frontal Gyrus |
| | | | | 7.4 | Precentral Gyrus |
| Decrease | 9 | 20 | 0.038 | 23.1 | Left Thalamus |
| | | | | 12.0 | Right Thalamus |
| Decrease | 10 | 21 | 0.046 | 33.3 | Posterior Cingulate Gyrus |
| | | | | 12.3 | Lingual Gyrus |
| | | | | 10.2 | Right Hippocampus |
| Decrease | 11 | 1261 | 0.003 | 18.6 | Right Putamen |
| | | | | 12.7 | Planum Temporale |
| | | | | 8.0 | Parietal Operculum Cortex |
| | | | | 7.6 | Heschl's Gyrus (includes H1 and H2) |
| Decrease | 12 | 2671 | 0.007 | 19.4 | Right Thalamus |
| | | | | 10.7 | Left Thalamus |
| | | | | 9.9 | Brain-Stem |
| | | | | 5.4 | Temporal Occipital Fusiform Cortex |
| Decrease | 13 | 6974 | <0.001 | 17.4 | Precentral Gyrus |
| | | | | 11.9 | Postcentral Gyrus |
| | | | | 6.7 | Anterior Cingulate Gyrus |
| | | | | 6.1 | Juxtapositional Lobule Cortex |
| *$D_1$-enriched FC* | | | | | |
| Increase | 8 | 13 | 0.034 | 60.0 | Inferior Lateral Occipital Cortex |
| | | | | 9.8 | Occipital Fusiform Gyrus |
| Increase | 9 | 26 | 0.042 | 23.6 | Postcentral Gyrus |
| | | | | 17.1 | Superior Parietal Lobule |
| Increase | 10 | 216 | 0.018 | 23.8 | Precuneus Cortex |
| | | | | 23.4 | Precentral Gyrus |
| | | | | 13.0 | Postcentral Gyrus |
| Increase | 11 | 232 | 0.008 | 34.9 | Precentral Gyrus |
| | | | | 24.9 | Postcentral Gyrus |
| Increase | 12 | 294 | 0.011 | 55.0 | Inferior Lateral Occipital Cortex |
| | | | | 11.3 | Middle Temporal Gyrus |
| Increase | 13 | 519 | 0.008 | 35.9 | Postcentral Gyrus |
| | | | | 12.8 | Superior Parietal Lobule |
| | | | | 7.8 | Precentral Gyrus |
| Decrease | 1 | 25 | 0.03 | 59.2 | Posterior Cingulate Gyrus |
| | | | | 15.5 | Anterior Cingulate Gyrus |
| *$D_{2/3}$-enriched FC* | | | | | |
| Increase | 12 | 22 | 0.022 | 46.2 | Temporal Pole |
| | | | | 20.0 | Frontal Orbital Cortex |
| Increase | 13 | 45684 | <0.001 | 6.8 | Precentral Gyrus |
| | | | | 6.3 | Postcentral Gyrus |
| | | | | 6.0 | Superior Lateral Occipital Cortex |

**Table S4. Cluster information for the REACT models with motion as covariate.** The table shows significant positive/negative target-enriched FC (pre-challenge) and significant increases/decreases in target-enriched FC (response to methylphenidate-challenge). Cluster information was extracted using FSL's autoaq. Clusters of ≥10 voxels and ≥5% overlap are reported. DAT=dopamine transporter; FC=functional connectivity; NAT=noradrenaline transporter.

| **Pre-challenge** | | | | | |
|---|---|---|---|---|---|
| | **Cluster** | **# voxels** | **Max p-value** | **Overlap (%)** | **Region** |
| *DAT-enriched FC* | | | | | |
| Positive | 2 | 191 | 0.01 | 83.5 | Right Putamen |
| | | | | 5.9 | Right Pallidum |
| Negative | 17 | 16 | 0.012 | 34.6 | Precentral Gyrus |
| | | | | 30.7 | Postcentral Gyrus |
| Negative | 18 | 21 | 0.032 | 31.6 | Precentral Gyrus |
| | | | | 16.5 | Superior Frontal Gyrus |
| | | | | 8.9 | Middle Frontal Gyrus |
| Negative | 19 | 25 | 0.023 | 34.7 | Superior Frontal Gyrus |
| | | | | 16.6 | Juxtapositional Lobule Cortex |
| Negative | 20 | 26 | 0.032 | 43.4 | Middle Frontal Gyrus |
| | | | | 20.3 | Precentral Gyrus |
| Negative | 21 | 26 | 0.042 | 30.9 | Postcentral Gyrus |
| | | | | 25.3 | Precentral Gyrus |
| Negative | 22 | 27 | 0.023 | 25.2 | Precentral Gyrus |
| | | | | 23.1 | Superior Frontal Gyrus |
| Negative | 23 | 31 | 0.043 | 63.3 | Frontal Pole |
| Negative | 24 | 34 | 0.025 | 48.0 | Left Thalamus |
| Negative | 25 | 59 | 0.039 | 79.1 | Left Thalamus |
| Negative | 26 | 81 | 0.033 | 38.4 | Planum Temporale |
| | | | | 13.7 | Central Opercular Cortex |
| | | | | 10.1 | Parietal Operculum Cortex |
| | | | | 7.6 | Heschl's Gyrus (includes H1 and H2) |
| Negative | 27 | 113 | 0.001 | 34.1 | Postcentral Gyrus |
| | | | | 27.0 | Precentral Gyrus |
| Negative | 28 | 220 | 0.02 | 29.8 | Middle Frontal Gyrus |
| | | | | 17.7 | Superior Frontal Gyrus |
| Negative | 29 | 265 | 0.028 | 32.6 | Frontal Pole |
| | | | | 31.7 | Frontal Orbital Cortex |
| Negative | 30 | 2886 | 0.01 | 25.2 | Frontal Pole |
| | | | | 14.6 | Middle Frontal Gyrus |
| | | | | 8.0 | Superior Frontal Gyrus |
| Negative | 31 | 28644 | <0.001 | 13.1 | Superior Lateral Occipital Cortex |
| | | | | 8.1 | Precuneus Cortex |
| | | | | 5.3 | Inferior Lateral Occipital Cortex |
| *NAT-enriched FC* | | | | | |
| Positive | 7 | 60 | 0.016 | 42.0 | Precentral Gyrus |
| | | | | 8.3 | Superior Frontal Gyrus |
| Positive | 8 | 98 | 0.003 | 29.7 | Precentral Gyrus |
| | | | | 10.7 | Superior Frontal Gyrus |
| | | | | 7.4 | Postcentral Gyrus |
| | | | | 5.1 | Juxtapositional Lobule Cortex |
| Positive | 9 | 782 | <0.001 | 41.7 | Left Thalamus |
| | | | | 29.4 | Right Thalamus |
| Positive | 10 | 2221 | <0.001 | 17.8 | Postcentral Gyrus |
| | | | | 10.0 | Precentral Gyrus |
| | | | | 7.4 | Central Opercular Cortex |
| | | | | 7.4 | Parietal Operculum Cortex |
| | | | | 6.4 | Planum Temporale |
| Positive | 11 | 2366 | <0.001 | 20.5 | Juxtapositional Lobule Cortex |
| | | | | 17.3 | Anterior Cingulate Gyrus |
| | | | | 14.8 | Precentral Gyrus |
| | | | | 6.4 | Paracingulate Gyrus |
| Positive | 12 | 2802 | <0.001 | 17.0 | Postcentral Gyrus |
| | | | | 13.7 | Central Opercular Cortex |
| | | | | 10.4 | Precentral Gyrus |

| | Cluster | # voxels | Max p-value | Overlap (%) | Region |
|---|---|---|---|---|---|
| | | | | 7.5 | Insular Cortex |
| | | | | 7.2 | Parietal Operculum Cortex |
| Negative | 1 | 15 | 0.035 | 40.0 | Inferior Lateral Occipital Cortex |
| | | | | 6.9 | Superior Lateral Occipital Cortex |
| *$D_1$-enriched FC* | | | | | |
| Positive | 3 | 88 | 0.024 | 40.6 | Frontal Orbital Cortex |
| | | | | 12.9 | Temporal Pole |
| | | | | 6.2 | Frontal Pole |
| Positive | 4 | 162 | 0.01 | 61.6 | Superior Lateral Occipital Cortex |
| | | | | 12.4 | Angular Gyrus |
| Positive | 5 | 952 | <0.001 | 46.6 | Posterior Cingulate Gyrus |
| | | | | 30.9 | Precuneus Cortex |
| | | | | 6.0 | Anterior Cingulate Gyrus |
| Positive | 6 | 3988 | <0.001 | 15.7 | Right Putamen |
| | | | | 14.6 | Left Putamen |
| | | | | 6.8 | Right Caudate |
| | | | | 6.7 | Left Caudate |
| Positive | 7 | 4454 | <0.001 | 23.4 | Paracingulate Gyrus |
| | | | | 20.1 | Frontal Pole |
| | | | | 13.3 | Anterior Cingulate Gyrus |
| | | | | 7.2 | Superior Frontal Gyrus |
| | | | | 5.7 | Middle Frontal Gyrus |
| Negative | 4 | 147 | 0.002 | 31.1 | Precentral Gyrus |
| | | | | 29.1 | Postcentral Gyrus |
| Negative | 5 | 16727 | <0.001 | 17.0 | Superior Lateral Occipital Cortex |
| | | | | 7.9 | Inferior Lateral Occipital Cortex |
| | | | | 5.1 | Occipital Pole |
| *$D_{2/3}$-enriched FC* | | | | | |
| Positive | 1 | 62 | 0.006 | 39.5 | Left Thalamus |
| | | | | 10.9 | Right Thalamus |
| Positive | 2 | 1401 | 0.001 | 28.2 | Right Putamen |
| | | | | 19.8 | Left Putamen |
| | | | | 8.4 | Right Caudate |
| | | | | 7.8 | Left Caudate |
| Negative | 4 | 10 | 0.008 | 47.9 | Superior Frontal Gyrus |
| Negative | 5 | 11 | 0.041 | 51.0 | Posterior Temporal Fusiform Cortex |
| | | | | 28.6 | Posterior Parahippocampal Gyrus |
| Negative | 6 | 23 | 0.037 | 34.3 | Left Thalamus |
| | | | | 15.5 | Left Hippocampus |
| Negative | 7 | 30270 | <0.001 | 10.3 | Superior Lateral Occipital Cortex |
| | | | | 5.6 | Precuneus Cortex |
| | | | | 5.4 | Inferior Lateral Occipital Cortex |

**Response to methylphenidate-challenge**

| | Cluster | # voxels | Max p-value | Overlap (%) | Region |
|---|---|---|---|---|---|
| *DAT-enriched FC* | | | | | |
| Increase | 8 | 10 | 0.034 | 26.5 | Inferior Frontal Gyrus |
| | | | | 18.9 | Inferior Frontal Gyrus |
| | | | | 10.3 | Middle Frontal Gyrus |
| Increase | 9 | 11 | 0.04 | 46.3 | Frontal Pole |
| Increase | 10 | 24 | 0.019 | 34.9 | Insular Cortex |
| | | | | 13.7 | Parietal Operculum Cortex |
| | | | | 11.6 | Central Opercular Cortex |
| Increase | 11 | 35 | 0.039 | 28.5 | Middle Temporal Gyrus |
| | | | | 18.7 | Posterior Superior Temporal Gyrus |
| | | | | 11.6 | Posterior Middle Temporal Gyrus |
| | | | | 9.2 | Posterior Supramarginal Gyrus |
| Increase | 12 | 50 | 0.034 | 51.3 | Temporal Pole |
| | | | | 10.1 | Anterior Middle Temporal Gyrus |
| | | | | 6.4 | Anterior Superior Temporal Gyrus |
| Increase | 13 | 86 | 0.026 | 37.6 | Precentral Gyrus |
| | | | | 18.8 | Inferior Frontal Gyrus |
| | | | | 10.8 | Middle Frontal Gyrus |
| Increase | 14 | 22026 | 0.001 | 5.5 | Postcentral Gyrus |

| | | | | | |
|---|---|---|---|---|---|
| | | | | 5.4 | Precentral Gyrus |
| | | | | 5.3 | Inferior Lateral Occipital Cortex |
| | | | | 5.2 | Superior Lateral Occipital Cortex |
| *NAT-enriched FC* | | | | | |
| Decrease | 5 | 23 | 0.04 | 38.9 | Heschl's Gyrus (includes H1 and H2) |
| | | | | 17.7 | Central Opercular Cortex |
| | | | | 9.9 | Planum Temporale |
| | | | | 5.7 | Planum Polare |
| Decrease | 6 | 48 | 0.031 | 45.6 | Superior Frontal Gyrus |
| Decrease | 7 | 102 | 0.015 | 36.7 | Parietal Operculum Cortex |
| | | | | 26.1 | Central Opercular Cortex |
| | | | | 11.6 | Heschl's Gyrus (includes H1 and H2) |
| Decrease | 8 | 367 | 0.02 | 34.8 | Precentral Gyrus |
| | | | | 33.2 | Postcentral Gyrus |
| Decrease | 9 | 3132 | <0.001 | 18.8 | Anterior Cingulate Gyrus |
| | | | | 17.4 | Precentral Gyrus |
| | | | | 9.2 | Juxtapositional Lobule Cortex |
| | | | | 8.1 | Postcentral Gyrus |
| | | | | 5.2 | Posterior Cingulate Gyrus |
| *$D_{2/3}$-enriched FC* | | | | | |
| Increase | 8 | 23 | 0.043 | 73.1 | Left Hippocampus |
| | | | | 6.1 | Anterior Parahippocampal Gyrus |
| Increase | 9 | 41 | 0.04 | 82.8 | Left Amygdala |
| Increase | 10 | 49 | 0.037 | 35.8 | Posterior Temporal Fusiform Cortex |
| | | | | 22.7 | Posterior Parahippocampal Gyrus |
| | | | | 8.2 | Left Hippocampus |
| | | | | 7.3 | Anterior Parahippocampal Gyrus |
| Increase | 11 | 162 | 0.022 | 31.1 | Frontal Pole |
| | | | | 29.2 | Inferior Frontal Gyrus |
| | | | | 9.1 | Middle Frontal Gyrus |
| Increase | 12 | 33510 | <0.001 | 7.0 | Precentral Gyrus |
| | | | | 6.4 | Postcentral Gyrus |
| | | | | 5.5 | Superior Lateral Occipital Cortex |

**Table S5. Cluster information showing significant associations between target-enriched FC and response inhibition, for the models without covariates.** The table shows significant positive/negative associations between pre-challenge response inhibition (dprime) and target-enriched FC, after Bonferroni-correction for multiple comparisons (4 maps tested, significance threshold $P_{FWE}$<0.0125). Cluster information was extracted using FSL's autoaq. Clusters of ≥10 voxels and ≥5% overlap are reported. FC=functional connectivity; NAT=noradrenaline transporter.

| **Pre-challenge** | | | | | |
|---|---|---|---|---|---|
| | **Cluster** | **# voxels** | **Max p-value** | **Overlap (%)** | **Region** |
| *NAT-enriched FC* | | | | | |
| Negative | 5 | 22 | 0.011 | 49.3 | Posterior Superior Temporal Gyrus |
| | | | | 22.0 | Posterior Middle Temporal Gyrus |
| Negative | 6 | 30 | 0.011 | 42.3 | Postcentral Gyrus |
| | | | | 16.4 | Precuneus Cortex |
| | | | | 8.7 | Precentral Gyrus |
| Negative | 7 | 31 | 0.011 | 46.9 | Precentral Gyrus |
| Negative | 8 | 50 | 0.01 | 42.5 | Parietal Operculum Cortex |
| | | | | 17.6 | Planum Temporale |
| | | | | 8.2 | Posterior Supramarginal Gyrus |
| Negative | 9 | 88 | 0.011 | 19.4 | Central Opercular Cortex |
| | | | | 17.6 | Planum Temporale |
| | | | | 12.3 | Parietal Operculum Cortex |
| | | | | 11.3 | Postcentral Gyrus |
| | | | | 6.9 | Posterior Superior Temporal Gyrus |
| | | | | 6.2 | Anterior Supramarginal Gyrus |
| Negative | 10 | 123 | 0.009 | 47.0 | Postcentral Gyrus |
| | | | | 7.6 | Precentral Gyrus |
| Negative | 11 | 374 | 0.006 | 40.3 | Postcentral Gyrus |
| | | | | 5.3 | Superior Parietal Lobule |
| *$D_1$-enriched FC* | | | | | |
| Negative | 2 | 18 | 0.008 | 47.6 | Subcallosal Cortex |
| *$D_{2/3}$-enriched FC* | | | | | |
| Positive | 1 | 42 | 0.007 | 24.2 | Superior Parietal Lobule |
| | | | | 13.3 | Precuneus Cortex |
| | | | | 7.7 | Superior Lateral Occipital Cortex |

**Supplementary Table S6. Spearman correlations of motion with clinical and cognitive measures.** Significant associations are shown in bold.

| | Total sample | Children | Adults |
|---|---|---|---|
| Response inhibition task performance (dprime) | | | |
| *Pre-challenge* | **R=-.57, P<.0001** | R=-.41, P=.09 | **R=-.39, P=.007** |
| *Response to challenge* | R=-.20, P=.11 | R=-.16, P=.49 | R=-.25, P=.10 |
| ADHD symptom severity - inattentive* | | | |
| *Pre-challenge* | R=.18, P=.15 | R=-.01, P=.98 | R=-.21, P=.17 |
| *Response to challenge* | R=-.09, P=.48 | R=-.09, P=.69 | **R=.34, P=.028** |
| ADHD symptom severity - hyperactive/impulsive* | | | |
| *Pre-challenge* | R=-.11, P=.38 | R=-.06, P=.78 | R=-.14, P=.35 |
| *Response to challenge* | R=-.05, P=.71 | R=-.13, P=.53 | R=.08, P=.60 |

*ADHD symptom severity sum scores were rescaled between 0 and 10 to facilitate comparability across age groups (for details, see Supplementary Methods).
